# WHO-Recommended Exercise Dose Is Associated With Slower Epigenetic Aging in Older Adults

**DOI:** 10.64898/2026.09.02.26361502

**Authors:** Ferenc Torma, Matyas Jokai, Gergely Babszky, Zsofia Babszky, Gabor Farkas, Soroosh Mozaffaritabar, Lei Zhou, Erika Koltai, Zsuzsanna Kneffel, Gabor Pavlik, Irina Kalabiska, Robert Olek, Eszter Petra Juranyi, Noémi Szántó, Peter Nagy, Bálazs Ligeti, Eszter Zita Martonné Szatmári, Csaba Kerepesi, Hajnalka Vágó, Robert T. Brooke, Steve Horvath, Zsolt Radak

## Abstract

The dose-response of six months of controlled exercise was studied in 72 subjects aged 50 to 70 years, with 1-4 training sessions per week, focusing on DNA methylation-based aging. Epigenetic age reduction increased with exercise dose, with about 255 MET-hours aligning with a one-year decrease in Robust PhenoAge acceleration - a volume consistent with WHO physical activity guidelines. The finding that relative ventricular posterior wall thickness and muscular quotient change in tandem with Robust PhenoAge acceleration suggests that epigenetic clocks reflect genuine organ-level rejuvenation. The DNA methylation-based protein prediction identified TGF-α, TIMP1, and adrenomedullin as showing a dose-response relationship with exercise volume. The exercise intervention yielded systemic health benefits, improved some cardiac parameters measured by echocardiogram, increased HDL levels, lowered blood pressure, reduced android fat mass, and notably, enhanced physiological functions, including cognitive performance. The program increased the relative abundance of microbes supporting intestinal barrier integrity and decreased pro-inflammatory bacterial populations. Serum N-lactoyl-phenylalanine levels showed a downward trend after six months of exercise, while trimethylamine N-oxide levels rose in male participants. This single-arm exercise study identified a dose–response relationship between cumulative exercise dose and Robust PhenoAge or GrimAge acceleration, strengthening the evidence that the observed associations are attributable to exercise exposure rather than background factors or confounders. The results further support the idea that regular physical activity can influence biological age by improving cardiac structure and function, and indicate that PhenoAge and GrimAge effectively reflect age-related cardiac changes.

## Introduction

It has been reported that occupational and leisure-time physical activity can reduce mortality rates ^1–3^, a finding that is particularly important for healthy aging. Stensvold et al. ^4^ examined the effects of a controlled five-year exercise intervention with high-intensity interval training (HIIT) and moderate-intensity continuous training (MICT) in 1567 healthy subjects (70-77 years). The study suggested that a combined MICT and HIIT intervention did not reduce all-cause mortality compared with recommended physical activity levels. However, a trend toward lower all-cause mortality was observed in the HIIT group compared with both the MICT and control groups ^4^. The HIIT group had a higher VO2max (0.7 mL/kg/min higher after the experimental period), which may be associated with a lower mortality risk, as a 1 mL/kg/min increase in VO2max has been associated with a reduced mortality rate^5^.

When the DNA methylation-based aging acceleration of healthy subjects with different levels of VO2max was compared, higher levels of physical fitness were associated with a deceleration of the estimated biological aging process ^6^ and Olympic Champions also showed a decreased rate of aging assessed by DNA methylation-based aging clocks ^7^. There are only few longitudinal controlled exercise intervention studies; one showed that eighteen women (50-70 yrs) participated in 8 weeks of combined exercise training. There was no change in epigenetic age acceleration in the treatment group; however, the exercise intervention reduced age acceleration in the subgroup with higher baseline acceleration ^8^. From an exercise physiology perspective, chronic adaptations arise from repeated training stimuli, the magnitude of which is determined by the cumulative exercise dose integrating exercise intensity, duration, frequency and baseline status. While current physical activity recommendations prescribe target exercise volumes, the cumulative exercise dose required to elicit favorable changes in DNA methylation-based biomarkers of aging has not been established.

Exercise-related adaptation is systemic and involves epigenetics and DNA methylation in this regulation ^7,9^, and is influenced by interorgan crosstalk via extracellular vesicles ^10^, the microbiome ^11^, and other molecular signaling pathways, ultimately contributing to improved physiological reserve.

Here, we conducted a prospective six-month supervised exercise intervention to investigate the dose-response relationship between cumulative exercise exposure and DNA methylation-based biological aging, while also evaluating physiological and cardiovascular function, body composition, and gut microbiome composition.

## Results

### Changes during the intervention period and associations with cumulative exercise dose

Higher MET-hour exposure was associated with a greater decrease (more favorable change) in epigenetic age acceleration marker Robust Pheno and Robust Grim-Age (Fig 1 A-B). Figure 1 panels C and D show model-derived estimates of the additional MET-hours required per 1-year decrease in epigenetic age acceleration for each epigenetic marker, based on the unadjusted model and the model adjusted for biological sex and chronological age, respectively. According to the unadjusted model, a one-year reduction in Robust PhenoAge acceleration corresponds to approximately 109.7 MET-hrs (sex- and age-adjusted model: 83.5 MET-hrs) over a six-month period. In the case of Robust GrimAge acceleration, an estimated 202 MET-hrs (sex- and age-adjusted model: 185.3 MET-hrs) per six months is required. The positive model intercept indicates that, at zero physical activity, the predicted age acceleration is above zero. Thus, in the absence of physical activity, positive age acceleration is expected. Based on the unadjusted model, approximately 255 MET-hours for Robust PhenoAge acceleration and 378 MET-hours for Robust GrimAge acceleration over six months would be required for an individual with no physical activity to achieve the one-year deceleration (i.e., to reach the exact - 1 year mark in the age acceleration axis if he or she is physically inactive). Notably, this magnitude of physical activity was broadly comparable to current WHO physical activity recommendations, corresponding to approximately a 260-520 MET-hrs range over the 6-month intervention period ^12^.

**Figure 1.**
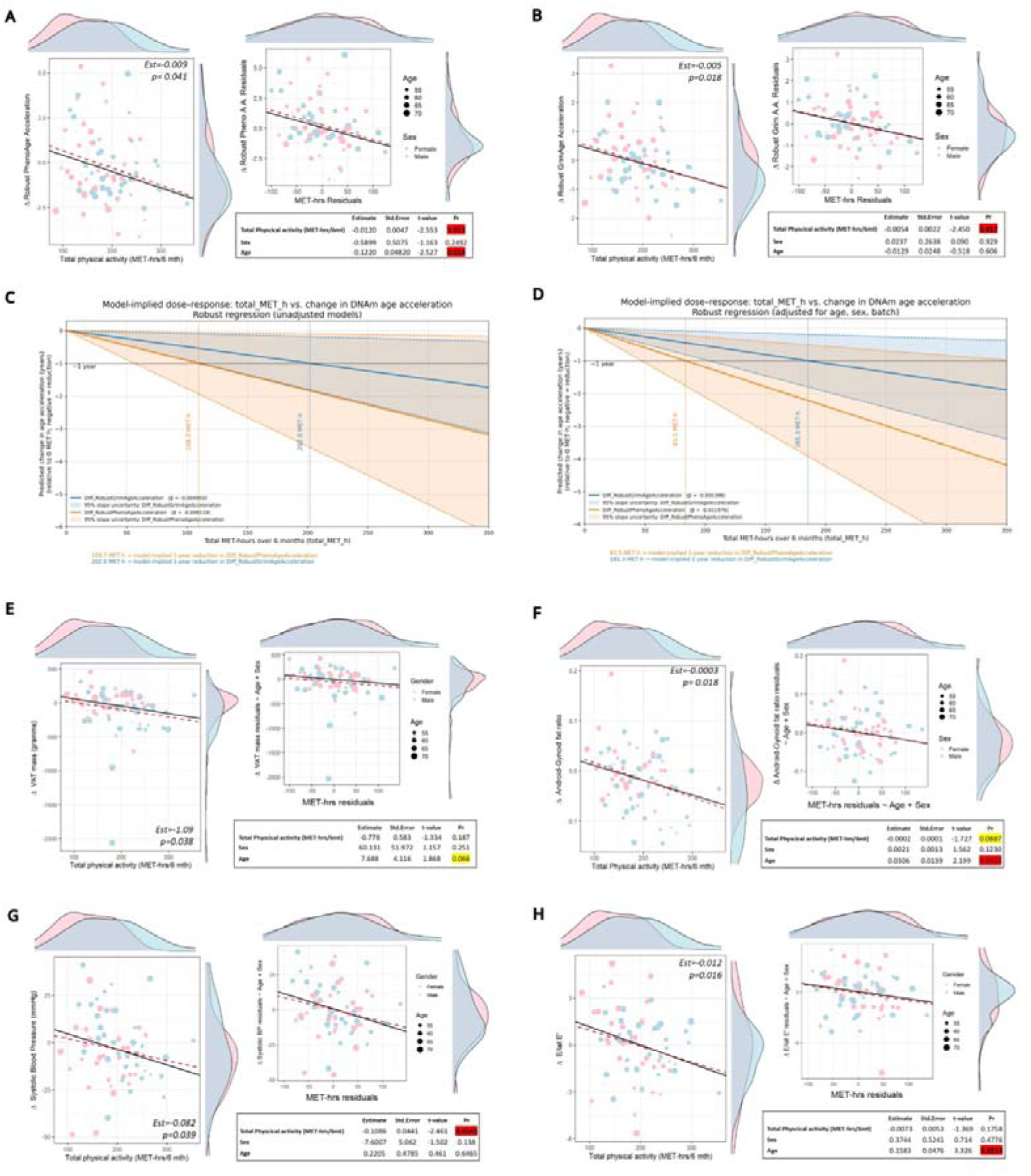
Robust associations between epigenetic aging indicators, physiological markers, and exercise dose. Panels A and B depict robust associations between epigenetic ageing markers and physical activity levels, expressed as MET-hours accumulated during the 6-month intervention period. Left panels show unadjusted robust regression models, while right panels present models adjusted for age and biological sex; corresponding statistical parameters are reported below each plot. Panels C and D illustrate, based on the unadjusted (C) and adjusted (D) models, the estimated level of physical activity required to reduce epigenetic age acceleration by one year for the respective epigenetic marker. Panels E–H show significant robust associations with physiological parameters. Pink markers represent female participants and light blue markers represent male participants. Solid lines indicate robust regression fits, whereas dashed red lines represent ordinary linear regression models. VAT: Visceral adipose tissue, E/letE′: Early transmitral flow to lateral mitral annular velocity ratio; a marker of left ventricular diastolic dysfunction.

Greater MET-hr levels are associated with greater loss of DEXA-measured visceral fat in the unadjusted robust model (Fig. 1 E) as well as decreased android to gynoid fat mass ratio (Fig. 1 F).

During the intervention, DNA methylation-related aging was reduced in all participants when DNAmFitAge Acceleration, Robust PhenoAge Acceleration, and Robust Hannum Age Acceleration were considered. Methylation-based telomere length decreased, and DunedinPACE is increased (Fig. 2 A). Gender-specific changes are shown in the panels in Fig. 2B and 2C.

**Figure 2.**
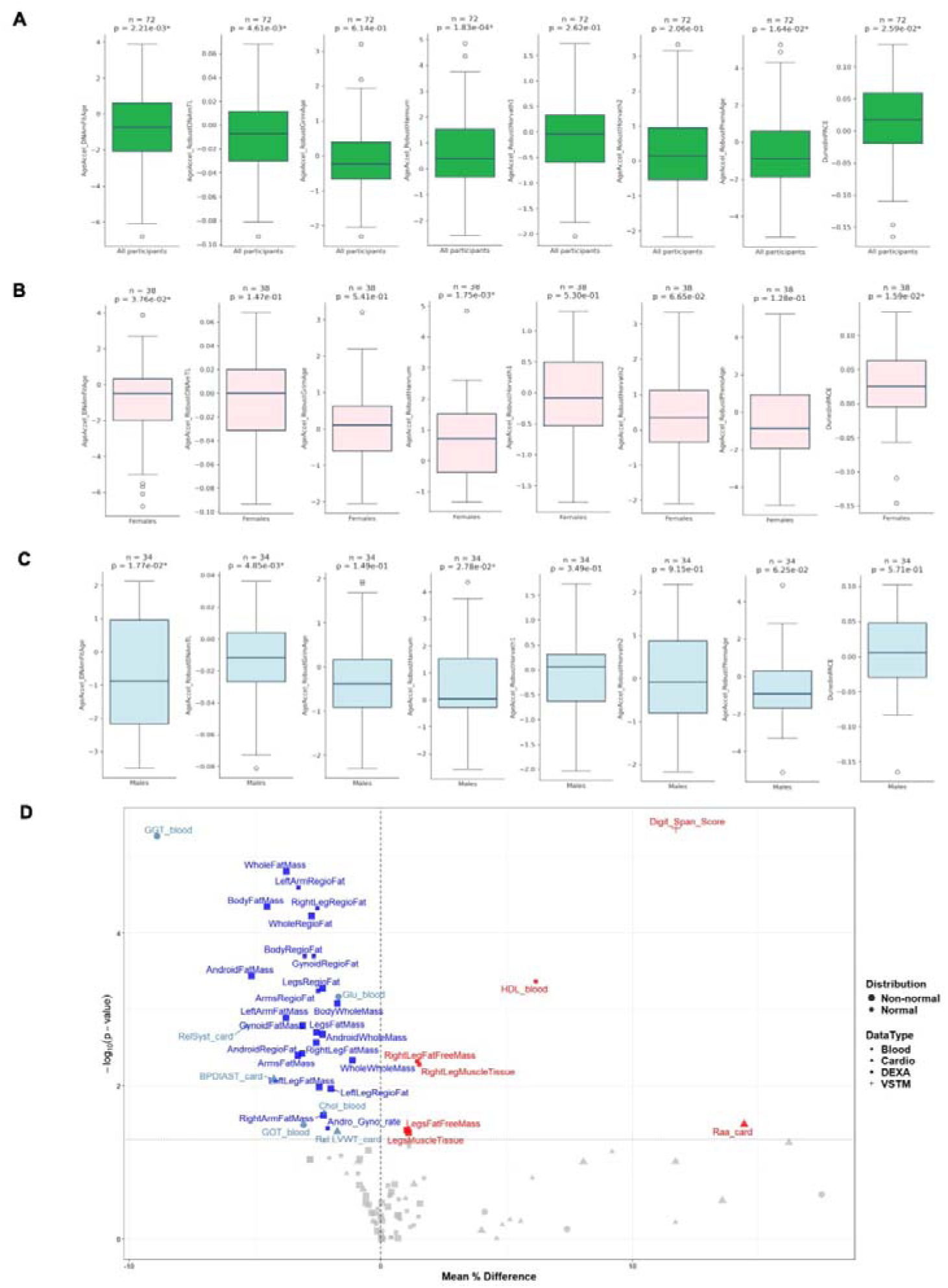
Epigenetic aging and physiological changes following a 6-month exercise intervention. Panel A shows changes in epigenetic clock acceleration in all participants, panel B in females only, and panel C in males only. Panel D presents before–after changes in the measured physiological indicators. For normally distributed data, paired t-tests were applied; when normality assumptions were violated, paired Wilcoxon signed-rank tests were used. Values are expressed as mean differences relative to baseline. In panels A–C, asterisks indicate statistically significant changes. In panel D, blue and light blue markers denote significant decreases, whereas red markers indicate significant increases.

DEXA results revealed decreases in body mass and fat mass and increased leg muscle tissue mass, along with several other anthropometric changes (Fig. 2 D). Circulating HDL levels increased during the intervention period, while the diastolic blood pressure decreased along with fasting serum glucose and total cholesterol (Fig. 2 D). Short term verbal memory was also improved indicated by the digit span test results (Fig. 2 D.) and the change in this cognitive test had reciprocal connection with methlyation based biological ageing indicated by the change in RobustGrimAge acceleration (Fig. 3 F).

**Figure 3.**
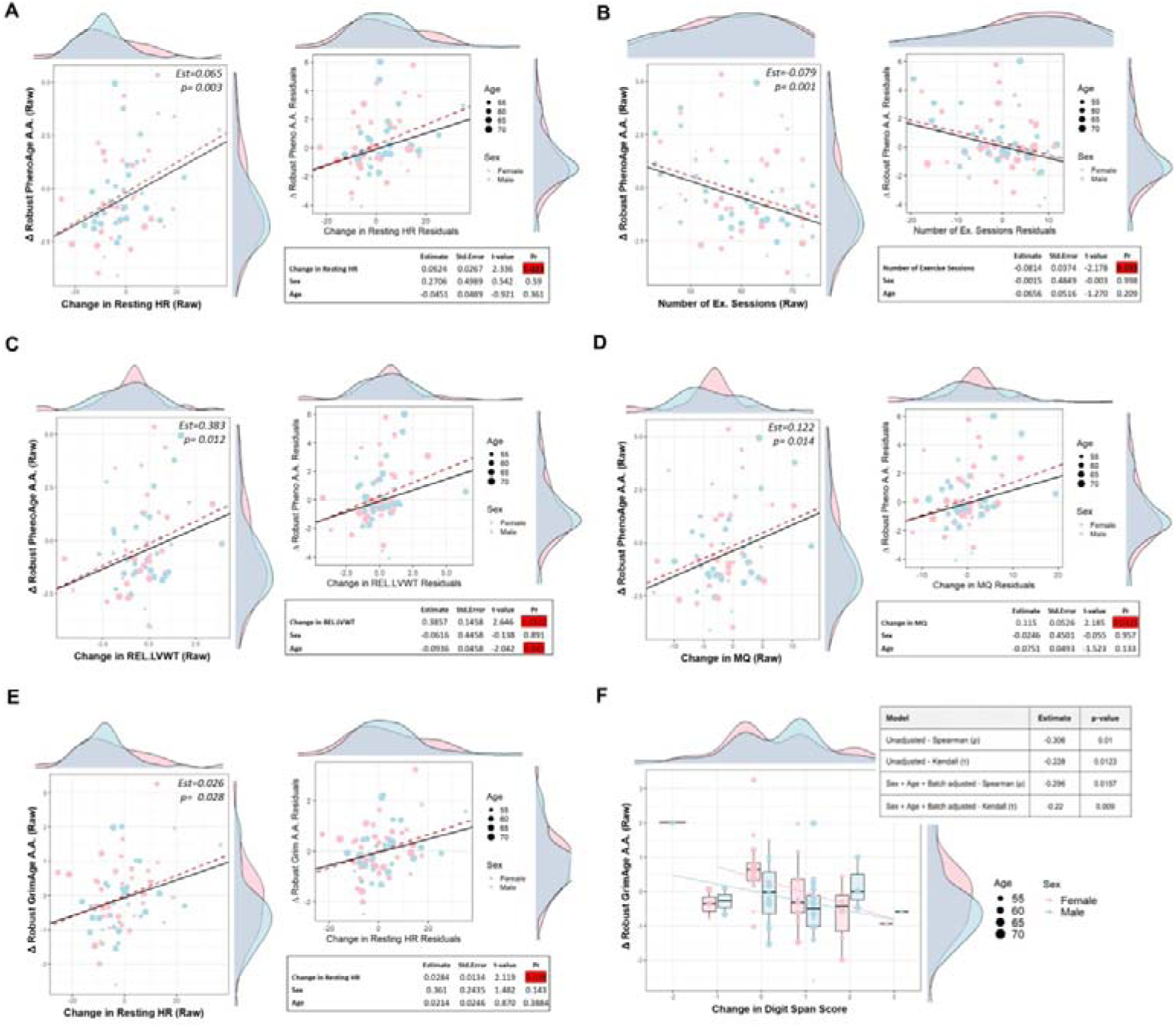
Robust associations between changes in epigenetic aging indicators and changes in exercise physiology and cardiovascular measures. Panels A–F show associations between the change in epigenetic age indicators (y-axis) and the change in selected physiological/cardiovascular variables (x-axis) across the intervention period. The epigenetic outcomes are Robust PhenoAge acceleration (A–D) and Robust GrimAge acceleration (E– F). The x-axis variables are change in resting heart rate (A, E), number of exercise sessions attended (B), REL.LVWT (C) and MQ (D). For panels A–E, left plots display unadjusted robust regression models, whereas right plots display robust regression models adjusted for chronological age and biological sex; corresponding effect estimates and p-values are reported below each plot. Panel F shows a nonparametric association between change in digit span score and change in Robust GrimAge acceleration. Points are colored by sex (pink: female; light blue: male). Solid lines indicate robust regression fits; dashed red lines indicate ordinary least-squares linear regression fits. REL.LVWT: wall thickness index defined as the sum of interventricular septal thickness (IVST) and left ventricular posterior wall thickness (LVPWT), normalized to the square root of body surface area (BSA), MQ ( Muscular Quotient): ratio of wall thickness to left ventricular end-diastolic diameter (EDD). Higher MQ values are indicative of a more concentric hypertrophic geometry.

The intervention resulted in beneficial changes in estimated VO2max and resting heart rate, as well as 1-min heart rate recovery, but maximal grip force remained unchanged at the group level (Figure S2).

### Echocardiographic changes and their relationship with cumulative MET-hours

In our cohort, higher total MET-hrs were negatively associated with the change in systolic blood pressure (SBP) in both the unadjusted and adjusted models, indicating that greater exercise volume was linked to a larger reduction in SBP (Fig. 1 G). In addition, in the unadjusted model, higher total MET-hrs were negatively associated with E/latE′ (the ratio of early transmitral inflow velocity to lateral mitral annular early diastolic velocity), indicating lower E/latE′ values with increasing exercise exposure (Fig. 1H). Because E/e′ is used as a non-invasive index of left ventricular filling pressures - with higher values suggesting higher filling pressures - the observed decrease in E/latE′ is consistent with a more favorable diastolic filling profile in participants with higher exercise volume. After the intervention period, Relative Systolic Fraction (RelSyst), an indicator describing systolic time relative to the whole cardiac cycle, showed a decrease - a shift that is typically characteristic of improved cardiovascular fitness (Fig2 C). Rel.LVWT also decreased at the group level. This is a left ventricular wall thickness marker, calculated as the sum of interventricular septal thickness (IVST) and left ventricular posterior wall thickness (LVPWT), normalized to the square root of body surface area (√BSA). In older, largely sedentary individuals, increased left ventricular wall thickness commonly reflects age- and inactivity-related concentric remodeling. The reduction observed following the intervention may indicates a reversal of this maladaptive remodeling, consistent with improved myocardial efficiency and reduced ventricular after-load.

### Links between epigenetic aging, exercise dose, and cardiac remodeling

Both Robust PhenoAge and Robust GrimAge acceleration showed robust associations with resting heart rate, a commonly used general fitness biomarker (Fig. 3 A and E). Robust PhenoAge acceleration was associated with the number of exercise sessions attended (Fig. 3 B), whereas Robust GrimAge acceleration was not; however, it was associated with total exercise volume expressed in MET-hours. This pattern suggests that cumulative exercise dose may be more relevant to biological age-related outcomes than simple session counts, highlighting the importance of intensity- and duration-weighted exposure metrics.

The change in Robust PhenoAge acceleration showed a positive robust linear association with the difference of Rel.LVWT and MQ in unadjusted and biological sex- and age-adjusted models ( Fig. 3 C and D). MQ is muscular quotient, defined as the ratio of left ventricular wall thickness to internal chamber diameter, and reflects left ventricular geometric remodeling. Higher values indicate a predominance of concentric hypertrophy, characterized by increased wall thickness relative to cavity size. Changes in these markers were positively associated with the change in aging rate indicated by Robust PhenoAge Acceleration. Notably, Rel.LVWT decreased significantly following the exercise intervention, whereas the muscular quotient showed only a trend toward reduction, not reaching statistical significance (p = 0.052).

### Associations with Methylation-Based Protein Estimates in Circulation

DNA methylation-based protein prediction reveals a hierarchy of exercise sensitivity among epigenetic biomarkers. TGF-α (Table S1) and TIMP1 (Table S2) showed the strongest robust negative associations with exercise dose after adjustment for age and sex, while adrenomedullin was associated with exercise dose only in the unadjusted analysis (Table S2).

Transforming growth factor-alpha (TGF-α) is a member of the epidermal growth factor family and regulates cell proliferation, survival, and circadian rhythm. It is induced by a single bout of exercise, while chronic exercise suppresses circulating TGF levels ^13^. Interestingly, at the protein level, TGF-α is positively associated with both Robust PhenoAge (Fig S3 B) and Robust GrimAge (Fig S3 C) acceleration,

We investigated whether changes in these surrogate markers explained any of the variation in echocardiographic parameters or blood biomarkers. Changes in Robust ADM showed significant positive associations with changes MQ in both unadjusted and age- and sex-adjusted robust regression analyses. Resting HR change showed a significant positive association with ADM change adjusted for sex and age. On the other hand change in E/A ratio showed a significant negative association with Robust ADM cahnge in the unadjusted robust analysis; however, this association was attenuated after adjustment for age and sex (Table S2). Among blood biomarkers, LDL cholesterol showed significant negative associations with Robust ADM change in both nonparametric and unadjusted robust analyses, whereas HDL cholesterol differnce showed a significant positive association in the unadjusted analysis.

Robust TIMP1 changes showed a consistent negative association with changes in LatE/A and a positive association with resting HR (Table S2). Additionally, the TIMP1 change showed a positive, nonparametric, and unadjusted robust association with Rel.LVWT and only positive nonparametric correlation with MQ, without a corresponding robust association, and only robust positive association with serum GGT levels. Overall, these predicted methylation-related proteins show a dose-response relationship with exercise load and may serve as potential biomarkers of adaptability to exercise training.

#### Microbial changes and Lac-Phe, TMAO related associations

Figure 4 presents the results of the shotgun metagenomic microbiome analysis, which is known to be strongly influenced by biological sex ^11^. Panels A-I represent the shift in different taxonomical units in a sex-stratified and overall manner. Similarly panel J-L shows bacterial pathway representations. At the genus level, Shannon diversity increased in males, females, and in the overall cohort (Fig. 4 M-O).

**Figure 4.**
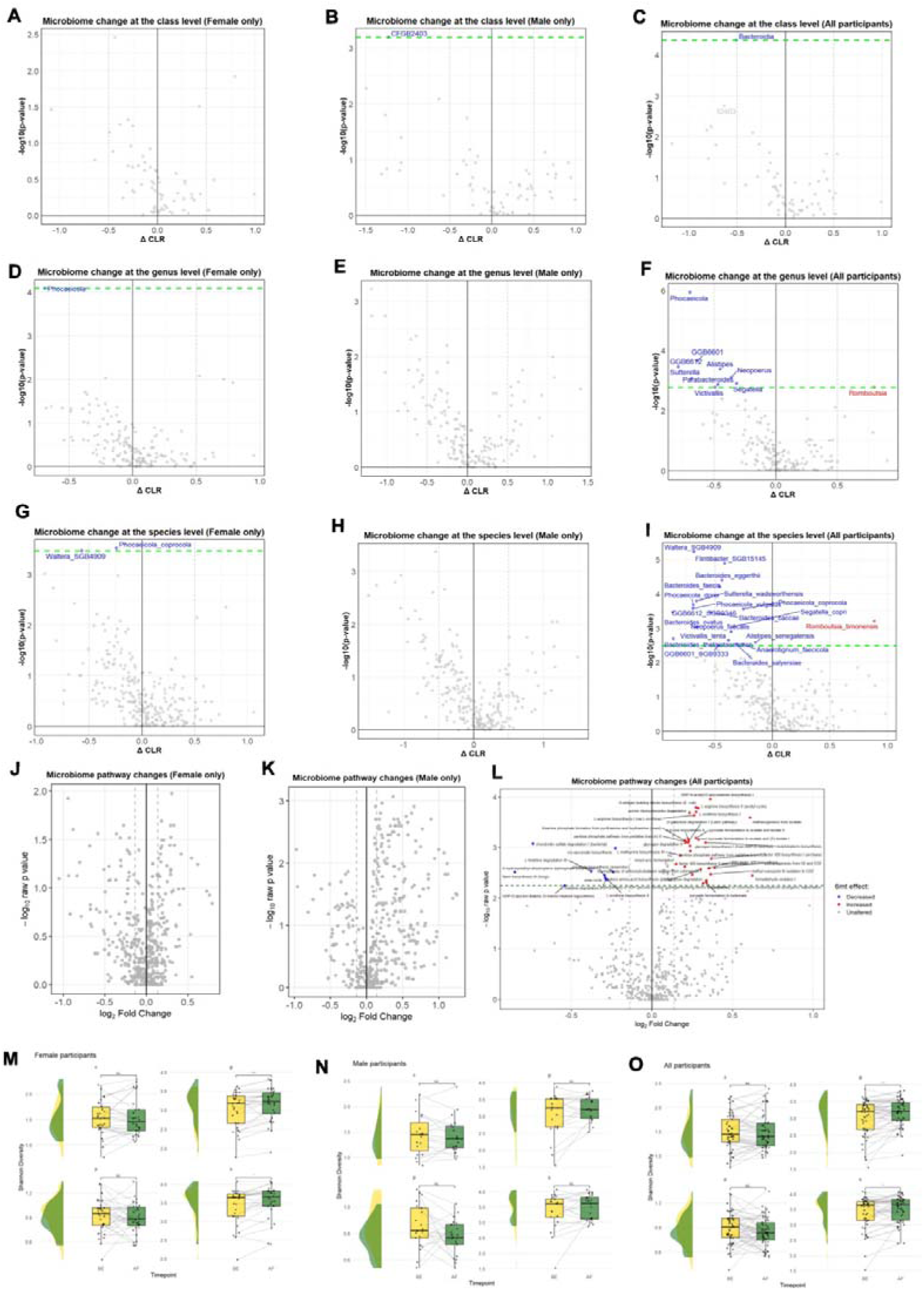
Changes in the microbiome during the intervention period. Panels A–I show CLR-transformed abundance changes during the intervention in females (A, D, G), males (B, E, H), and all participants (C, F, I) at the class (A–C), genus (D–F), and species (G–I) levels. Panels J–L depict microbiome pathway changes in females, males, and all participants, respectively. Panels N–P show Shannon diversity values before (BE) and after (AF) the intervention. Taxonomic abbreviations are as follows: c, class; p, phylum; g, genus; s, species. In the volcano plots, blue dots indicate significantly decreased abundance, red dots indicate significantly increased abundance, and gray dots indicate non-significant changes. The green dashed line represents the FDR significance threshold. NS, not significant; *p < 0.05; **p < 0.01; ***p < 0.001.

We detected increased UDP-N-acetyl-D-glucosamine biosynthesis I (Fig. 4L and Table S3), which may be important for maintaining gut integrity and resistance to inflammation ^14^. We found that during the six-month training, the production of Tryptophan was increased along with Tyrosine, Phenylalanine, L-ornithine biosynthesis I, and L-arginine biosynthesis I in the gut, which is overall beneficial for the host organism by maintaining gut barrier integrity, modulating the immune and neurotransmitter systems, and decreasing cardiovascular risk ^15^. The training-related increase in purine ribonucleoside degradation by the microbiome could mean that proper regulation of purine levels decreases the risk of gout. The increased pyruvate fermentation to acetate and lactate, and (S)-lactate, could be important to short-chain fatty acid (SCFA) production and cross-feeding of microbes. The exercise-induced increases in microbial sucrose biosynthesis II could mean carbon storage, osmoprotection, and enhanced metabolic flexibility.

TMAO is a product of gut bacterial metabolism of choline, L-carnitine, betaine, and phosphatidylcholine, which are abundant in red meat, fish, and other seafood ^16^. Results are conflicting regarding the health-related role of TMAO levels; some studies report health-threatening effects ^17^, while others report beneficial effects ^18^. Here, we found an increase in male participants following the exercise intervention (Fig. S4 A).

N-lactoyl-phenylalanine, a peptide conjugate of lactate and phenylalanine created from (S)-lactate and L-phenylalanine by the cytosol nonspecific dipeptidase (CNDP2) protein ^19^. Lac-Phe, found to be an exercise-induced metabolite that controls appetite ^20^, might be involved in the regulation of inflammation ^21^ and aging ^22^. Here, we report that serum Lac-Phe levels showed a decreasing trend after six months of exercise training (Wilcoxon test, p = 0.08; Fig. S4B); the change in older subjects was greater than in younger subjects (Fig. S4C).

## Discussion

DNA methylation-based aging clocks use a selected set of cytosines that correlate with chronological age to evaluate aging progression, mortality, and susceptibility to various diseases as personalized biomarkers ^23^. Robust PhenoAge and Robust GrimAge acceleration were associated with MET-hours, indicating that even relatively modest levels of physical activity may contribute to improved biological aging trajectories and reduced disease risk. Elite swimmers, or Tour de France cyclists, for example, could have about 250-400 MET hours per week, representing a workload approximately 29-47 times higher than that estimated by our most conservative model to achieve an additional one-year reduction in epigenetic age acceleration. The dose-response relationship between MET and age acceleration is unlikely to be linear; nevertheless, a given increase in MET may meaningfully reduce the rate of biological aging. Indeed, we found that Olympic champions’ (aged between 25 and 102 years) epigenetic age is about 6 years lower than their chronological age; those who were closer to their active sport career showed greater benefit than those who finished decades ago ^7^. Tour de France cyclists also showed an increase in average longevity (17%) when compared with the general population ^24^. Our data suggest that regular exercise, through DNA methylation, regulates gene expression and longevity.

The exercise intervention provided all systemic benefits, indicated by our echocardiogram-measured cardiac parameters, increased HDL levels, decreased blood pressure, and, importantly, improved physiological functions, including cognitive function, assessed by the digit span test. We also observed a decrease in android fat mass during the exercise program, consistent with a previous report showing that six months of exercise training alters DNA methylation in human adipose tissue, potentially influencing adipocyte metabolism and contributing to reductions in fat mass ^25^.

It is well known that exercise-induced systemic adaptation includes changes in the gut microbiome ^26^, and a link has been reported between DNA methylation age acceleration and the microbiome ^11^. Here, we show that a six-month exercise program increased the relative abundance of microbes that support the integrity of the intestinal barrier and reduced pro-inflammatory bacterial populations, which we found to slow epigenetic aging ^11^.

Here, we observed an increase in TMAO levels in male participants after the exercise intervention. TMAO has been identified as a gut flora-derived agent that causes and predicts CVD ^27^, cerebral infarction ^28^, and promotes age-related endothelial dysfunction via oxidative stress ^29^. On the other hand, in a rat model of advanced heart failure, chronic TMAO administration significantly improved survival and was accompanied by favorable hemodynamic changes, including reduced arterial pressure ^30^. Additionally, evidence from a clinical study in hemodialysis patients showed that L-carnitine supplementation-induced TMAO elevation was associated with reduced markers of vascular injury and oxidative stress ^31^. TMAO has been shown to enhance blood-brain barrier integrity, protect against inflammatory insults, and improve cognitive function in mice. These effects are accompanied by reduced reactivity in astrocytes and microglia ^32^. Thirteen-day TMAO administration markedly prolonged exhaustive swimming time in mice and increased levels of 3-hydroxybutyrate, isocitrate, anserine, TMA, taurine, glycine, and glutathione, and disrupted three metabolic pathways related to oxidative stress and protein synthesis in skeletal muscle ^33^. It has also been reported that TMAO levels decrease after exercise and rebound to baseline levels ^34^. Data suggest that exercise alters TMAO levels ^35^, but the mechanisms remain unclear.

Here, we report that serum Lac-Phe levels showed a decreasing trend after 6 months of exercise training. The change was more pronounced in older subjects compared to younger ones. Acute exercise has been shown to increase circulating Lac-Phe levels in an intensity-dependent manner ^36^; however, the effects of long-term exercise training on Lac-Phe remain largely unknown. Lac-Phe has been identified as an appetite-suppressing agent; high-intensity exercise and Lac-Phe administration decreased appetite and weight in mice ^20^. The impact of prolonged exercise training on circulating Lac-Phe in humans remains poorly understood.

Aging is associated with cardiac muscle remodeling, as age is related to left ventricular hypertrophy in elderly subjects, even after controlling for 24hr BP ^37^. This hypertrophy is distinct from the “athlete heart,” since age-related hypertrophy is characterized by increased deposition of fat, collagen, and lipofuscin in the myocardium ^38^ and is accompanied by impaired diastolic function and reduced cardiac reserve ^39^.

RelSyst and Rel.LVWT decreased during the intervention phase, accompanied by reductions in diastolic blood pressure and resting heart rate, and improvements in heart rate recovery. In addition, changes in SBP were negatively associated with exercise volume (MET-hours) in both adjusted and unadjusted models, while E/latE’ showed a negative association with exercise volume only in the unadjusted model. Taken together, these findings may reflect favorable cardiovascular adaptations to 6-month exercise, potentially involving both structural and autonomic mechanisms. The reduction in RelSyst may reflect a relative shortening of systolic time and a more favorable redistribution of the cardiac cycle toward diastole, a pattern generally consistent with improved ventricular filling and potentially better cardiac efficiency ^40,41^. Similarly, the decrease in Rel.LVWT may suggest subtle remodeling toward a less concentric phenotype; however, this interpretation should be made with caution. Given that Rel.LVWT is indexed to body surface area; changes in body composition should be taken into account when interpreting this parameter. However, reduced body fat alone would not readily explain a lower Rel.LVWT, as a decrease in BSA would tend to increase the indexed value. In this context, prior interventional studies have shown that prolonged exercise training can reduce LV stiffness and increase LV end-diastolic volume in previously sedentary middle-aged adults, broadly consistent with a favorable remodeling response ^42^.

The observed reductions in resting heart rate and improvements in heart rate recovery are consistent with enhanced parasympathetic tone and/or reduced sympathetic activity, well-established adaptations to regular exercise training ^43^. The association between exercise volume and reductions in SBP further supports dose-dependent peripheral adaptations, potentially including improved endothelial function ^44^ and reduced arterial stiffness ^45^. Although the negative association between E/latE’ and exercise volume did not remain significant after adjustment, it may tentatively suggest improvements in diastolic function or filling pressures. However, given the lack of robustness, this finding should be interpreted as exploratory.

With regard to biological aging, MQ and Rel. LVWT was associated with higher Robust PhenoAge acceleration, even after adjustment for age and sex. As both indices reflect cardiac geometry and hypertrophic remodeling, these findings may indicate that second-generation epigenetic clocks are sensitive to aspects of cardiovascular remodeling beyond chronological age and biological sex. However, this interpretation remains speculative and requires confirmation in future studies.

In conclusion, six months of exercise training at WHO-recommended MET levels, in addition to conferring functional benefits to skeletal muscle, the circulatory system, the gut microbiome, and the brain, appears to be associated with a slowing of DNA methylation-based age acceleration. Importantly, these effects also appear to follow a dose-dependent pattern. The physiological significance of chronic exercise-induced increases in circulating TMAO and decreases in Lac-Phe requires further investigation.

## Methods

### Study Population and Design

A total of 78 participants (36 males, 42 females) were enrolled in this intervention study investigating the longitudinal effects of relatively moderate HIIT. Participants had no history of cardiovascular disease, pulmonary disease, psychiatric or neurological disorders, cancer, musculoskeletal disorders, or degenerative arthrosis, and none reported excessive smoking. All participants underwent physician screening to confirm eligibility for exercise. Cognitive status was assessed using the Montreal Cognitive Assessment (MoCA). All study protocols and procedures were conducted in accordance with the Declaration of Helsinki and were approved by the relevant local governmental ethics committee (NNGYK/14120-7/2025). Written informed consent was obtained from all participants prior to enrollment.

Participation was voluntary, with sessions offered three times per week. Adherence varied among the cohort, with total session attendance ranging between 15 and 74 sessions per participant.

Four participants dropped out due to personal circumstances or health-related issues, and two additional participants were excluded from analysis due to low attendance (<35 sessions). Physiological testing (VO2max estimation, grip force) was conducted throughout the intervention period, with assessments performed after every 6-8 exercise sessions. Blood samples were collected at both pre- and post-intervention time points from 74 participants. Dual-energy X-ray absorptiometry (DEXA), echocardiographic data, blood-based methylation biomarkers and total physical activity volume (MET-hrs) were available for 69–72 participants. Baseline descriptive statistics and average exercise attendance are summarized in Table 1.

**Table 1.** Participant Characteristics.

| Sex | N | Age (years) | BMI (kg/m<sup>2</sup>) | Est. VO<sub>2</sub>max (mL·kg<sup>-1</sup>·min<sup>-1</sup>) | Average session MET | Exercise session attendance |
| --- | --- | --- | --- | --- | --- | --- |
| Female | 38 | 59.9 ± 5.1 | 25.2 ± 4.2 | 36.1 ± 5.2 | 5.7 ± 1.0 | 59.9 ± 8.9 |
| Male | 34 | 62.1 ± 5.1 | 27.4 ± 3.7 | 42.3 ± 5.2 | 6.9 ± 1.2 | 59.7 ± 9.1 |

Sex denotes biological sex; Age represents chronological age at the beginning of the intervention period; Average session MET represents the mean exercise intensity achieved by each sex group throughout the intervention program. Data are presented as mean ± SD.

Microbiological samples were available from 52 participants at both baseline and post-intervention assessments.

### Exercise Intervention

The exercise intervention utilized a multi-modal approach, incorporating stationary bicycles, elliptical trainers, and indoor rowing ergometers. The initial phase of the program consisted of six introductory familiarization sessions designed to acclimate participants to the equipment and workload (Fig. S1 A). After the initial six sessions, participants underwent a relative HIIT intervention for six months, during which the training volume was increased over time (Fig. S1 B-D). Detalis of the protocols are presented in the extended methods section

### Limitations

The absence of a nonintervention control group limits causal interpretation of the observed pre-post changes. However, the primary aim was to examine dose-response associations between exercise exposure and DNA methylation-based aging within the intervention cohort rather than the efficacy of HIIT. Randomized controlled studies are needed to confirm these findings.

## Data Availability

All data produced in the present study are available upon reasonable request to the authors

## Author contributions

F.T., M.J., and Z.R. wrote the original draft of the manuscript. F.T., M.J., G.B., Z.B., G.F., S.M., and L.Z. contributed to data collection, data curation, and formal analysis. E.K., R.O., E.P.J., N.S., and P.N. performed the biochemical analyses. Z.K., G.P., and H.V. performed the cardiovascular assessments. I.K. performed the anthropometric and dual-energy X-ray absorptiometry analyses. B.L. performed the microbiome analyses. E.Z. M.S., C.K., R.T.B., S.H., and F.T. performed the bioinformatic and statistical analyses. Z.R., F.T., C.K., and P.N. edited the manuscript. Z.R. supervised the study. All authors reviewed and approved the final manuscript.

## Acknowledgments

The authors gratefully acknowledge the contribution of Dorina Annár to the DEXA body composition measurements. The authors declare no conflicts of interest relevant to this work.

## Data Sharing Statement

Full blood DNA methylation data generated during the current study are available in the Gene Expression Omnibus (GEO) repository under accession number: [GSEXXXXXX]. Other data are available from the corresponding author upon reasonable request.

## References

1 Morris, J. N., Heady, J. A., Raffle, P. A., Roberts, C. G. & Parks, J. W. Coronary heart-disease and physical activity of work. Lancet 262, 1111–1120; concl, doi:10.1016/s0140-6736(53)91495-0 (1953).

2 Gao, W., Sanna, M., Chen, Y. H., Tsai, M. K. & Wen, C. P. Occupational Sitting Time, Leisure Physical Activity, and All-Cause and Cardiovascular Disease Mortality. JAMA Netw Open 7, e2350680, doi:10.1001/jamanetworkopen.2023.50680 (2024).

3 Lear, S. A. et al. The effect of physical activity on mortality and cardiovascular disease in 130 000 people from 17 high-income, middle-income, and low-income countries: the PURE study. Lancet 390, 2643–2654, doi:10.1016/S0140-6736(17)31634-3 (2017).

4 Stensvold, D. et al. Effect of exercise training for five years on all cause mortality in older adults-the Generation 100 study: randomised controlled trial. BMJ 371, m3485, doi:10.1136/bmj.m3485 (2020).

5 Keteyian, S. J. et al. Peak aerobic capacity predicts prognosis in patients with coronary heart disease. Am Heart J 156, 292–300, doi:10.1016/j.ahj.2008.03.017 (2008).

6 Jokai, M. et al. DNA methylation clock DNAmFitAge shows regular exercise is associated with slower aging and systemic adaptation. GeroScience 45, 2805–2817, doi:10.1007/s11357-023-00826-1 (2023).

7 Radak, Z. et al. Slowed epigenetic aging in Olympic champions compared to non-champions. GeroScience 47, 2555–2565, doi:10.1007/s11357-024-01440-5 (2025).

8 da Silva Rodrigues, G., et al. Eight Weeks of Physical Training Decreases 2 Years of DNA Methylation Age of Sedentary Women. Res Q Exerc Sport 95, 405–415, doi:10.1080/02701367.2023.2228388 (2024).

9 Radak, Z. et al. Epigenetic and “redoxogenetic” adaptation to physical exercise. Free Radic Biol Med 210, 65–74, doi:10.1016/j.freeradbiomed.2023.11.005 (2024).

10 Gyorgy, B. et al. The protein cargo of extracellular vesicles correlates with the epigenetic aging clock of exercise sensitive DNAmFitAge. Biogerontology 26, 35, doi:10.1007/s10522-024-10177-9 (2025).

11 Torma, F. et al. Alterations of the gut microbiome are associated with epigenetic age acceleration and physical fitness. Aging cell 23, e14101, doi:10.1111/acel.14101 (2024).

12 Bull, F. C. et al. World Health Organization 2020 guidelines on physical activity and sedentary behaviour. Br J Sports Med 54, 1451–1462, doi:10.1136/bjsports-2020-102955 (2020).

13 Costanti-Nascimento, A. C. et al. Physical exercise as a friend not a foe in acute kidney diseases through immune system modulation. Front Immunol 14, 1212163, doi:10.3389/fimmu.2023.1212163 (2023).

14 Wang, K. S. et al. Alterations of gut microbiome in chronic rhinosinusitis: insights from a mendelian randomization study. Braz J Otorhinolaryngol 92, 101698, doi:10.1016/j.bjorl.2025.101698 (2026).

15 Li, T. T. et al. Microbiota metabolism of intestinal amino acids impacts host nutrient homeostasis and physiology. Cell Host Microbe 32, 661–675 e610, doi:10.1016/j.chom.2024.04.004 (2024).

16 Koeth, R. A. et al. Intestinal microbiota metabolism of L-carnitine, a nutrient in red meat, promotes atherosclerosis. Nature medicine 19, 576–585, doi:10.1038/nm.3145 (2013).

17 Cameron, S. J. et al. Circulating Trimethylamine N-Oxide and Growth Rate of Abdominal Aortic Aneurysms and Surgical Risk. JAMA Cardiol 10, 1000–1009, doi:10.1001/jamacardio.2025.2698 (2025).

18 Chen, Y. L. et al. Endometrial stromal cell-derived TMAO sustains decidualization to prevent recurrent spontaneous abortion. Cell Metab 38, 316–330 e318, doi:10.1016/j.cmet.2025.11.014 (2026).

19 Jansen, R. S. et al. N-lactoyl-amino acids are ubiquitous metabolites that originate from CNDP2-mediated reverse proteolysis of lactate and amino acids. Proc Natl Acad Sci U S A 112, 6601–6606, doi:10.1073/pnas.1424638112 (2015).

20 Li, V. L. et al. An exercise-inducible metabolite that suppresses feeding and obesity. Nature 606, 785–790, doi:10.1038/s41586-022-04828-5 (2022).

21 Ying, W. et al. N-Lactoyl-Phenylalanine modulates lipid metabolism in microglia/macrophage via the AMPK-PGC1alpha-PPARgamma pathway to promote recovery in mice with spinal cord injury. J Neuroinflammation 22, 167, doi:10.1186/s12974-025-03495-3 (2025).

22 Liu, P. L. et al. Bilayer Scaffolds Synergize Immunomodulation and Rejuvenation via Layer-Specific Release of CK2.1 and the “Exercise Hormone” Lac-Phe for Enhanced Osteochondral Regeneration. Adv Healthc Mater 14, e2402329, doi:10.1002/adhm.202402329 (2025).

23 Moqri, M. et al. Validation of biomarkers of aging. Nature medicine 30, 360–372, doi:10.1038/s41591-023-02784-9 (2024).

24 Sanchis-Gomar, F., Olaso-Gonzalez, G., Corella, D., Gomez-Cabrera, M. C. & Vina, J. Increased average longevity among the “Tour de France” cyclists. Int J Sports Med 32, 644–647, doi:10.1055/s-0031-1271711 (2011).

25 Ronn, T. et al. A six months exercise intervention influences the genome-wide DNA methylation pattern in human adipose tissue. PLoS Genet 9, e1003572, doi:10.1371/journal.pgen.1003572 (2013).

26 Tseng, C. H. & Wu, C. Y. From dysbiosis to longevity: a narrative review into the gut microbiome’s impact on aging. J Biomed Sci 32, 93, doi:10.1186/s12929-025-01179-x (2025).

27 Wang, Z. et al. Gut flora metabolism of phosphatidylcholine promotes cardiovascular disease. Nature 472, 57–63, doi:10.1038/nature09922 (2011).

28 Wang, L., Nan, Y., Zhu, W. & Wang, S. Effect of TMAO on the incidence and prognosis of cerebral infarction: a systematic review and meta-analysis. Front Neurol 14, 1287928, doi:10.3389/fneur.2023.1287928 (2023).

29 Brunt, V. E. et al. Trimethylamine-N-Oxide Promotes Age-Related Vascular Oxidative Stress and Endothelial Dysfunction in Mice and Healthy Humans. Hypertension 76, 101–112, doi:10.1161/HYPERTENSIONAHA.120.14759 (2020).

30 Gawrys-Kopczynska, M. et al. TMAO, a seafood-derived molecule, produces diuresis and reduces mortality in heart failure rats. Elife 9, doi:10.7554/eLife.57028 (2020).

31 Fukami, K. et al. Oral L-carnitine supplementation increases trimethylamine-N-oxide but reduces markers of vascular injury in hemodialysis patients. J Cardiovasc Pharmacol 65, 289–295, doi:10.1097/FJC.0000000000000197 (2015).

32 Hoyles, L. et al. Regulation of blood-brain barrier integrity by microbiome-associated methylamines and cognition by trimethylamine N-oxide. Microbiome 9, 235, doi:10.1186/s40168-021-01181-z (2021).

33 Zou, H. et al. Effects of Trimethylamine N-Oxide in Improving Exercise Performance in Mice: A (1)H-NMR-Based Metabolomic Analysis Approach. Molecules 29, doi:10.3390/molecules29174128 (2024).

34 Pechlivanis, A. et al. Monitoring the Response of the Human Urinary Metabolome to Brief Maximal Exercise by a Combination of RP-UPLC-MS and (1)H NMR Spectroscopy. J Proteome Res 14, 4610–4622, doi:10.1021/acs.jproteome.5b00470 (2015).

35 Miccheli, A. et al. The influence of a sports drink on the postexercise metabolism of elite athletes as investigated by NMR-based metabolomics. J Am Coll Nutr 28, 553–564, doi:10.1080/07315724.2009.10719787 (2009).

36 Weber, D., Ferrario, P. G. & Bub, A. Exercise intensity determines circulating levels of Lac-Phe and other exerkines: a randomized crossover trial. Metabolomics 21, 63, doi:10.1007/s11306-025-02260-0 (2025).

37 Toba, A. et al. Impact of age on left ventricular geometry and diastolic function in elderly patients with treated hypertension. Blood Press 26, 264–271, doi:10.1080/08037051.2017.1306422 (2017).

38 Kitzman, D. W. & Edwards, W. D. Age-related changes in the anatomy of the normal human heart. J Gerontol 45, M33–39, doi:10.1093/geronj/45.2.m33 (1990).

39 Dai, D. F., Chen, T., Johnson, S. C., Szeto, H. & Rabinovitch, P. S. Cardiac aging: from molecular mechanisms to significance in human health and disease. Antioxid Redox Signal 16, 1492–1526, doi:10.1089/ars.2011.4179 (2012).

40 Nazario Leao, R., et al. Systolic time ratio measured by impedance cardiography accurately screens left ventricular diastolic dysfunction in patients with arterial hypertension. Clin Hypertens 23, 28, doi:10.1186/s40885-017-0084-y (2017).

41 Levy, W. C., Cerqueira, M. D., Abrass, I. B., Schwartz, R. S. & Stratton, J. R. Endurance exercise training augments diastolic filling at rest and during exercise in healthy young and older men. Circulation 88, 116–126, doi:10.1161/01.cir.88.1.116 (1993).

42 Howden, E. J. et al. Reversing the Cardiac Effects of Sedentary Aging in Middle Age-A Randomized Controlled Trial: Implications For Heart Failure Prevention. Circulation 137, 1549–1560, doi:10.1161/CIRCULATIONAHA.117.030617 (2018).

43 Imai, K. et al. Vagally mediated heart rate recovery after exercise is accelerated in athletes but blunted in patients with chronic heart failure. Journal of the American College of Cardiology 24, 1529–1535, doi:10.1016/0735-1097(94)90150-3 (1994).

44 Goto, C. et al. Effect of different intensities of exercise on endothelium-dependent vasodilation in humans: role of endothelium-dependent nitric oxide and oxidative stress. Circulation 108, 530–535, doi:10.1161/01.CIR.0000080893.55729.28 (2003).

45 Lopes, S. et al. Exercise training reduces arterial stiffness in adults with hypertension: a systematic review and meta-analysis. J Hypertens 39, 214–222, doi:10.1097/HJH.0000000000002619 (2021).

