## Supplementary information for "WHO-Recommended Exercise Dose Is Associated With Slower Epigenetic Aging in Older Adults"

**Tabble of content**

Extended Materials and Merthods	2

Supplementary Figures (S1–S12)	7

Supplementary Tables (S1–S3)	11

Supplementary References	28

**Extended Materials and Merthods**

#### **HIIT Exercise Protocol and Intensity Monitoring**

Cardiovascular intensity was monitored continuously during the exercise using Polar H10 heart rate sensors interfaced with the Polar Beat - Polar Flow software ecosystem. To establish target intensity zones, each participant's maximal heart rate (HRmax) was estimated using the Tanaka formula ^1^:

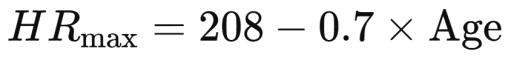

This calculation served as the physiological baseline for monitoring and regulating exercise intensity throughout the remainder of the six-month intervention.

After the initial 6 exercise sessions that had a duration of 34 minutes. For the subsequent sessions (7th through 25th), a standardized 34-minute protocol was implemented (supplementary figure 2 B). Each session commenced with a 12-minute warm-up phase, progressively increasing in intensity to ensure participants reached approximately 70% of their estimated maximal heart rate (HRmax) by the twelfth minute.

The high-intensity component of the session consisted of five intervals, each lasting two minutes. During these work bouts, participants were instructed to elevate their heart rate above 80% of their (HRmax). Each high-intensity interval was followed by a 2-minute active recovery period at a reduced intensity; participants were required to maintain movement throughout the recovery phase.

To ensure participant safety and adherence to the prescribed effort levels, intensity was monitored using a dual-method approach. In addition to heart rate tracking, the Borg Rating of Perceived Exertion (RPE) scale (0–10) was utilized. A target threshold was established to ensure that perceived exertion did not exceed a value of 8 on the Borg scale. Consequently, in specific instances where the subjective RPE reached the threshold of 8 before the physiological target was met, participants were permitted to maintain that level even if their heart rate remained below 80% of HRmax

Starting from the 26th session through the 45th, the exercise protocol was modified to increase the total volume of high-intensity work. The warm-up duration was reduced to 8 minutes, while the total session duration remained constant at 34 minutes. The 4 minutes redistributed from the warm-up phase allowed for the inclusion of an additional high-intensity interval. Consequently, the modified protocol required participants to perform six work bouts exceeding 80% of their HRmax, as opposed to the five intervals performed in the previous phase. Each work bout continued to be followed by a 2-minute active recovery period, maintaining the established work-to-rest ratio while increasing the overall physiological demand of the session.

From the 46th session onward, the protocol was further expanded to include an additional high-intensity interval and a subsequent recovery period. This adjustment involved the addition of one 2-minute work bout and one 2-minute active recovery phase at the end of the existing routine. Consequently, the total session duration was extended to 38 minutes. This final progression increased the total number of high-intensity intervals to seven per session, increasing the cumulative time spent above the 80% HRmax threshold while maintaining the established interval structure.

**Physiological and Functional Testing and VO₂max Estimation**

To assess autonomic cardiac function and track changes in physical fitness, heart rate data were recorded at specific rest intervals. At the commencement of each session, participants remained in a sedentary, resting position for 2 minutes to establish the pre-exercise resting heart rate (RHR) for that day. Furthermore, immediately following the final high-intensity interval of the session, participants returned to the same resting position. This allowed for the measurement of one-minute heart rate recovery (HRR). RHR, mean heart rate, estimated maximum heart rate, estimated VO₂max, and exercise duration were used to calculate total exercise volume, expressed as MET-hours. Details of the exercise intervention and calculations are provided in the Extended Methods section.

Relative aerobic capacity was monitored by estimating relative maximal oxygen uptake (VO_2_max) using the Chester Step Test (CST) protocol, as previously described ^2^. Total physical activity effort was assessed using MET-hours, calculated from heart rate-derived variables, estimated VO_2_max and exercise duration.

To monitor changes in overall systemic strength, handgrip strength (HGS) was measured using a Camry EH101 electronic handgrip dynamometer. This assessment served as a proxy for total body muscular strength.

Verbal short term memory was assessed by the digit span test ^3,4^.

### ***Epigenetic biomarkers of ageing***

DNA methylation–based biological age estimators were computed using the DNA Methylation Age Calculator provided by the Clock Foundation Team (<https://dnamage.clockfoundation.org/clock>). The following estimators were included: **Horvath’s Skin & Blood clock ^5^**, **Horvath’s pan-tissue clock ^5^**, **DNAm FitAge ^6^**, and blood-derived clocks including the **robust** implementations of **DNAm PhenoAge**, **GrimAge ^7,8^**, the **Hannum blood-based clock ^9^ and the DunedinPACE ^10^**. Circulating protein EpiScores were calculated as described previously ^11^.

In addition, a DNA methylation-based **telomere length estimator** was considered ^12^. Robust epigenetic clocks (e.g., Robust PhenoAge and Robust GrimAge) were implemented using the **MethylCIPHER** R package. These robust variants extend the original formulations by applying additional statistical procedures intended to improve reliability and reduce susceptibility to technical artifacts and biological heterogeneity ^13^.

**Biochemical analysis**

***Lac-Phe measurement***

20 ul of human serum samples were thawed, and 60 ul of 75% methanol was added. Samples were centrifuged at 14 000 rcf for 10 min at 4°C and the supernatant was acidified with 10% formic acid. Before the injection, samples were diluted to a two-fold dilution in 0.1% FA in water. Liquid chromatography-tandem mass spectrometry (LC-MS/MS) measurements were performed using a Thermo Q-Exactive Focus Orbitrap mass spectrometer linked to a Thermo Vanquish UHPLC system. The analysis was performed using a Phenomenex Kinetex C18 column (50 x 2.1 mm, 2.6 µm) with eluents 0.1% FA/H2O (A) and 0.1% FA/MeOH (B). The gradient started with 5% B, which increased linearly to 13% in 2 minutes, then to 95% in 4 minutes, held there for 0.5 minutes, then decreased back to 5% B in 0.1 minutes, and remained at that level for 3.4 minutes before the next injection. The flow rate was 0.5 ml/min, and the column was maintained at 40°C. Detection was conducted in negative ionization mode, with higher-energy collisional dissociation (HCD) employed to identify Lac-Phe (236>88).

***Determination of Trimethylamine N-oxide (TMAO)levels***

Plasma trimethylamine N-oxide (TMAO) concentrations were determined using liquid chromatography coupled with tandem mass spectrometry (LC–MS/MS). Briefly, 50 μL of plasma was mixed with 100 μL of a deuterated internal standard solution (TMAO-d9) and incubated at 36°C with shaking (1500 rpm) for 15 min. Subsequently, 600 μL of acetonitrile was added for protein precipitation, followed by incubation at 36°C with shaking (700 rpm) for 30 min. After centrifugation (3000 rpm, 10 min), 200 μL of the clear supernatant was transferred to a 96-well polypropylene plate for LC–MS/MS analysis.

Chromatographic separation was performed on a Kinetex HILIC column (100 Å, 50 × 2.1 mm, 2.6 μm; Phenomenex) maintained at 35°C, while the autosampler temperature was set to 8°C. The mobile phase consisted of (A) 0.2% acetic acid in water/acetonitrile (1:1, v/v) containing 10 mmol/L ammonium acetate and (B) acetonitrile. The flow rate was 0.65 mL/min, the injection volume was 10 μL, and the total run time was 6 min using a gradient elution program.

Mass spectrometric detection was performed on a QTRAP 3200 triple quadrupole mass spectrometer (AB Sciex) equipped with an electrospray ionization (ESI) source operating in positive ion mode and using multiple reaction monitoring (MRM). Quantification was based on isotope dilution using TMAO-d9 as the internal standard. Calibration curves were prepared in TMAO-free human serum over a concentration range of 10–2000 ng/mL. The concentration of TMAO in each sample was calculated from the analyte-to-internal standard peak area ratio using the calibration curve.

***Microbiome metagenome analysis***

The bioinformatic analysis followed a standard shotgun metagenomics workflow. In brief, the quality of paired-end sequencing reads was evaluated using FastQC v0.12.1 and MultiQC v1.31 ^14^. Low-quality reads were filtered and trimmed with fastp v1.0.1 ^15^. During preprocessing, paired-end adapter sequences and polyG/polyX tails were removed, low-quality bases at both read ends were trimmed using a Phred quality score cutoff of 20, and reads shorter than 50 bp were discarded. Reads originating from the host were eliminated by aligning against the human reference genome (GRCh38_noalt_as) with Bowtie2 v2.5.4 ^16^. Taxonomic profiling of host-filtered reads was conducted using MetaPhlAn v4.1.2 ^17^, while functional profiling was performed with HUMAnN v3.9 ^18^ using the UniRef90 protein database ^19^. The resulting taxonomic profiles, gene family abundances, and pathway abundance tables were subsequently used for downstream analyses.

### Taxa with a relative abundance below 0.005% or detected in fewer than 15% of samples were removed prior to downstream analyses. Alpha diversity was estimated using the Shannon diversity indices at each taxonomic level ^20^. For compositional data analysis, zeros were imputed using Bayesian-multiplicative replacement implemented in the R package zCompositions ^21,22^, followed by centered log-ratio (CLR) transformation ^23^ using the scikit-bio Python library ^24^. Finally, taxa with a median CLR value below -2 were excluded from subsequent statistical analyses.

### ***Statistical analysis***

All statistical analyses were performed in **R (v4.4.1)**, using the **ppcor**, **purrr**, and **robustbase** packages. Prior to inferential testing, variable distributions were assessed using the **Shapiro-Wilk test**. Because several variables deviated from normality, and to preserve potentially informative biological variation, primary association analyses were conducted using **robust linear regression** based on **MM-type estimators** for both unadjusted and covariate-adjusted models ^25,26^.

Shotgun metagenomic data were processed as described earlier ^27^. and processed using a standardized bioinformatics pipeline for quality control, host read removal, and taxonomic and functional profiling, followed by compositional data preprocessing and filtering.

Within-subject pre-post differences were tested using a **paired t-test** when normality assumptions were acceptable; otherwise, the **Wilcoxon signed-rank test** was applied. To evaluate whether changes differed from zero (e.g., slope of estimated VO2max, slope of maximal grip force), **one-sample t-tests** or **one-sample Wilcoxon signed-rank tests** were used, as appropriate. **Spearman’s rank correlation** was used for associations involving ordinal/discrete outcomes (e.g., Digit Span scores) and for data types where rank-based methods were preferable, including compositional microbiome measures.

***Total physical effort assessment (MET-hours)***

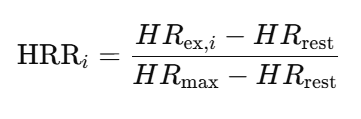
For calculating the MET-hours during the 6 month intervention period HR-data was monitore dduring each exercise session. Providing the average exercise intensity expressed in beats per minute. From the resting HR, average HR and the calculated HR-max hear rate reserve franction (HRR) was calculated was calculated for each exercise training:
 from the %HRR ≈ %VO_2_max association ^28^ MET cost was calculated for each exercise bouts. Total MET-hours were then calculated as the sum, across all exercise bouts (i...N), of heart-rate-reserve–scaled VO_2_max intensity multiplied by exercise duration (t_i_) and deviled by the Metabolic Equivalent.

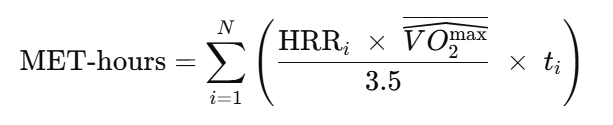

Where VO2max is considered as the individual average value of all the estimated values during the 6 month intervention period.

***Body composition assessment***

Two main components were reported for each participant in the DEXA mesurement: fat mass (FM) and fat-free mass (FFM). Fat mass represents the total weight of adipose tissue in the body and is expressed in grams and as a percentage of total body mass. Fat-free mass includes all non-fat tissues (skeletal muscle, bone mineral, organs, and residual soft tissues) and was also given in absolute values (g; kg) and as a percentage of body mass. Segmental DEXA analysis enabled regional differentiation of FM and FFM, including trunk, upper limbs, lower limbs, and android/gynoid regions, which is particularly useful for assessing body-composition adaptations to training.

We performed the DEXA total body measurement according to the manufacturer’s standard protocol, with participants lying in a supine position, arms at the sides, legs together, and minimal clothing, avoiding metallic objects. The scan typically lasted 8–15 minutes, depending on the selected body-coverage mode and the participant’s body mass. In heavier individuals, the measurement time was longer to allow additional scan passes or higher-definition settings, ensuring adequate data quality.

#### ***Biochemical Analysis and Blood Sampling (ext.)***

Venous blood samples were collected from each participant at baseline (pre-intervention) and post-intervention (no exercise 48 h before the sampling). The sampling protocol included three 3 mL EDTA tubes and one 8 mL serum-separating tube (SST). Following centrifugation, the plasma fractions were pipetted into Eppendorf tubes and stored for further analysis. After the collection, the samples were stored at -80°C.

Genomic DNA was isolated from EDTA-anticoagulated whole blood samples and normalized to a concentration of 50 ng/µl prior to DNA methylation analysis. Bisulfite conversion was performed using the EZ-96 DNA Methylation MagPrep Kit (Zymo Research, Irvine, CA, USA). Genome-wide DNA methylation profiling was then carried out using the Illumina Infinium MethylationEPIC BeadChip array, following the manufacturer’s standardized protocols, as previously described ^29^.

The SST samples were processed at room temperature via centrifugation at 3000 rpm for 10 minutes. The resulting serum was utilized for clinical blood chemistry analysis, including the quantification of the following metabolic and hepatic parameters: glucose (Glu), triglycerides (Tg), total cholesterol (Chol), high-density lipoprotein (HDL), low-density lipoprotein (LDL), glutamate-oxaloacetate transaminase (GOT), glutamate-pyruvate transaminase (GPT), and gamma-glutamyl transferase (GGT).

TGF-alpha concentration was measured from undiluted blood serum samples of the participants using an R&D Systems Immunoassay Kit (Cat. No. DTGA00). The protocol was performed according to the manufacturer's instructions using the sandwich-ELISA technique, and the optical density was detected with a CLARIOstar Plus microplate reader at 450 nm using wavelength correction.

##### **Echocardiographic Parameters**

Transthoracic echocardiography was performed to examine the potential cardiac remodeling effects of the training intervention. During the echocardiographic examination (Dornier AI 4800, Germany), the following parameters were recorded before and after the exercise program: VCF, LVMM_N, CON_N, E/A, LVET/QT, BPS, BPD, REL.LVWT, REL.LVID, MQ, E/medE', medS’, medE'/A', latE'/A', latS', RVDA, relsyst, E/latE', TAPSE, LAA, RAA, and LVMM as described below

Relative left ventricular muscle mass (rel.LVMM) was calculated as LVMM normalized to body surface area (BSA^3/2). Relative cardiac output (rel.CO) was defined as CO normalized to BSA^3/2.

Transmitral flow velocities included peak early diastolic filling velocity (E) and peak late (atrial) filling velocity (A), with their ratio (E/A) used as an index of diastolic function, where higher values indicate more favorable filling dynamics. The ratio of left ventricular ejection time to electrical systole (LVET/QT) reflects the relationship between mechanical and electrical systole.

Blood pressure variables included systolic blood pressure (BPSyst) and diastolic blood pressure (BPDiast).

Relative left ventricular wall thickness (Rel.LVWT) was calculated as LVWTd normalized to BSA^1/2, where LVWTd represents the sum of interventricular septal diastolic thickness and left ventricular posterior wall thickness in diastole. Relative left ventricular internal diameter (Rel.LVID) was defined as LVID normalized to BSA^1/2, where LVID corresponds to the left ventricular internal diameter in diastole.

The muscular quotient (MQ) was calculated as the ratio of combined left ventricular wall thickness to internal diameter (LVWTd/LVIDd).

Tissue Doppler imaging parameters included medial (septal) and lateral mitral annular velocities: early diastolic (medE’, latE’), late diastolic (medA’, latA’), and systolic velocities (medS’, latS’).

Filling pressure indices were assessed using E/medE’ and E/latE’, where lower values indicate more favorable left ventricular filling pressures.

Right ventricular and atrial measurements included right ventricular diastolic area (RVDA), tricuspid annular plane systolic excursion (TAPSE), left atrial area (LAA), and right atrial area (RAA). Relative systolic duration (RelSyst) was defined as the proportion of systole within the total cardiac cycle.

##### **Body Composition Parameters (DEXA)**

Body composition was evaluated using Dual-Energy X-ray Absorptiometry (DEXA, type: Lunar Prodigy, enCORE v18). This diagnostic tool provided several distinct parameters per scan, allowing for a comprehensive, segmental analysis of tissue distribution. The assessment provided precise data on adipose tissue (fat mass) and fat-free mass, which includes lean mass (skeletal muscle), organs, and mineralized tissue (bone mass), across various anatomical regions

Regional measurements were obtained for the arms, legs, trunk, android, and gynoid compartments, including right-left side-specific values and their differences. These included fat mass, lean tissue, fat-free mass, and total tissue mass.

Total body measures included total fat mass, total lean tissue, total fat-free mass, and total mass, with corresponding right, left, and difference values where applicable.

Relative composition was further characterized by regional fat percentage and tissue fat percentage across all compartments (arms, legs, trunk, android, gynoid), as well as total and side-specific distributions. Visceral adiposity was quantified using visceral fat tissue volume and mass.

**Supplementary Figures**

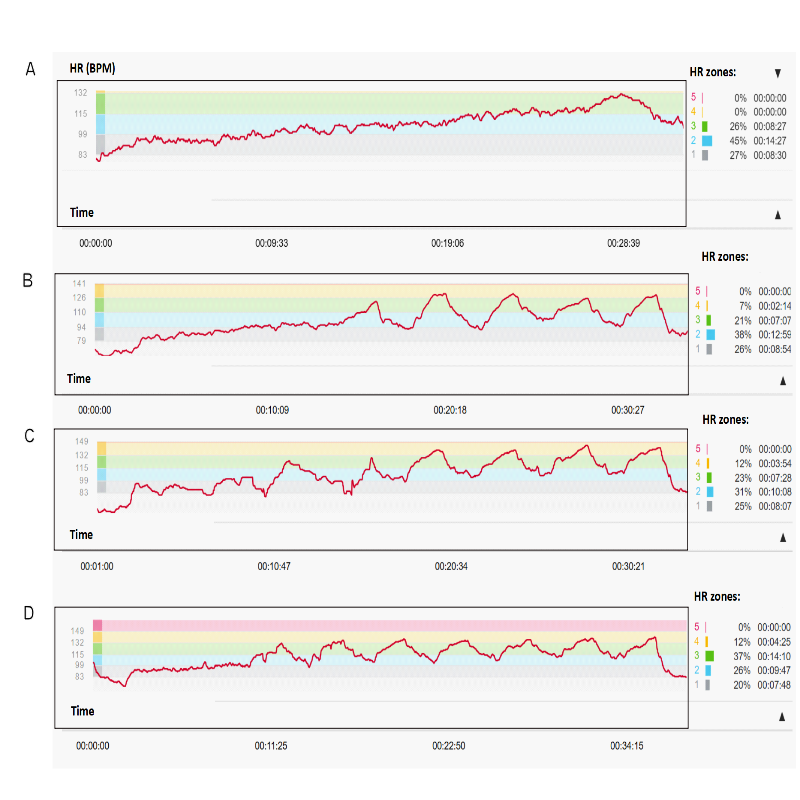

**Supplementary Figure 1. | Representative exercise sessions during the intervention program.**

Panel A represents sessions from the first 6 weeks, followed by panels depicting sessions from weeks 7–25 (B), sessions 26–45 (C), and finally sessions from the 46th session onward until the end of the intervention (D).

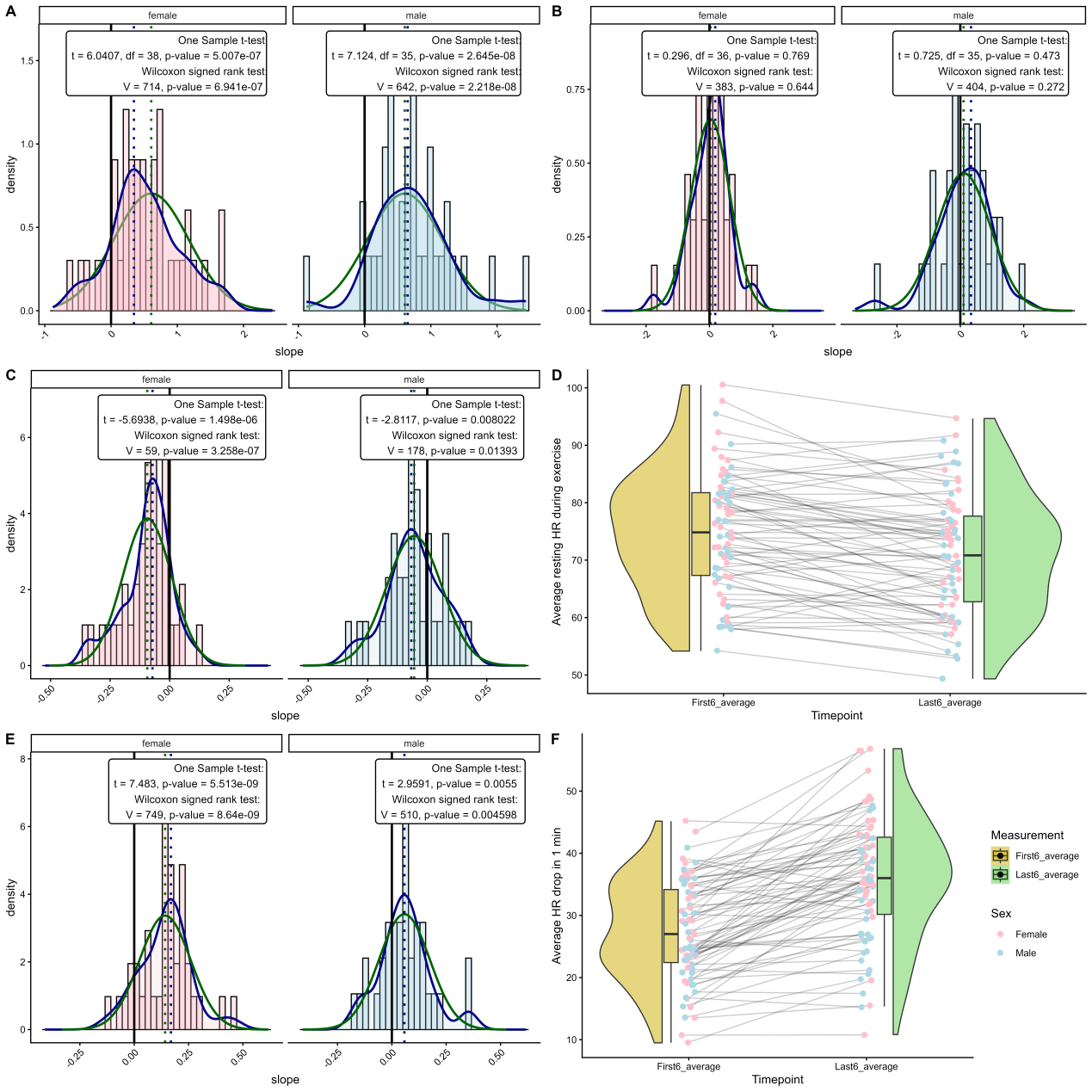

**Supplementary Figure 2. | Exercise physiology markers during the 6-month intervention period.**

Panel A presents the degree of change (slope) in estimated VO₂max, assessed using the Chester Step Test, during the intervention in female and male participants. Panel B shows trends in maximal handgrip strength. Panels C and D depict changes in resting heart rate during exercise, expressed as the slope across all measurement points (C) and as the average values of the first and last six sessions (D). Similarly, panels E and F show 1-minute heart rate recovery after exercise in the seated position, expressed as the slope across all measurements (E) and as the average values of the first and last six sessions (F). Density distributions are shown as histograms colored pink for females and blue for males. Parametric and non-parametric tests assessing deviation from zero are displayed above the histograms, as indicated. *: p<0.05.

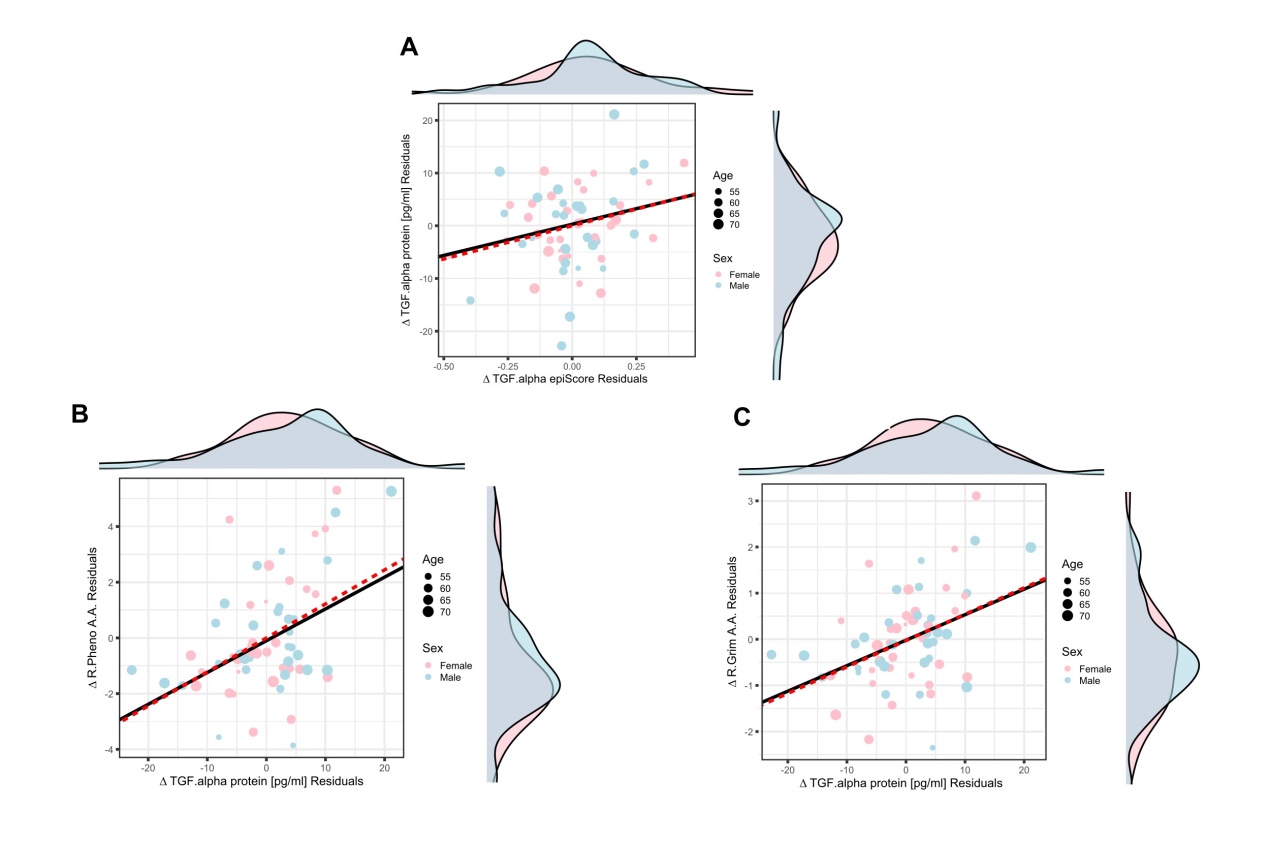

**Supplementary Figure 3. | TGF-ɑ protein assications with methylation based protein and ageing markers**

Linear association between serum TGF-ɑ protein measured by ELISA and epiScore change (A), and the cahnges in Robust PhenoAge (B) and RobustGrimAge (C) accelration. All linear models were adjusted for biological sex, chronological age and batch. Pink markers represent female participants and light blue markers represent male participants. Solid lines indicate robust regression fits, whereas dashed red lines represent ordinary linear regression models.

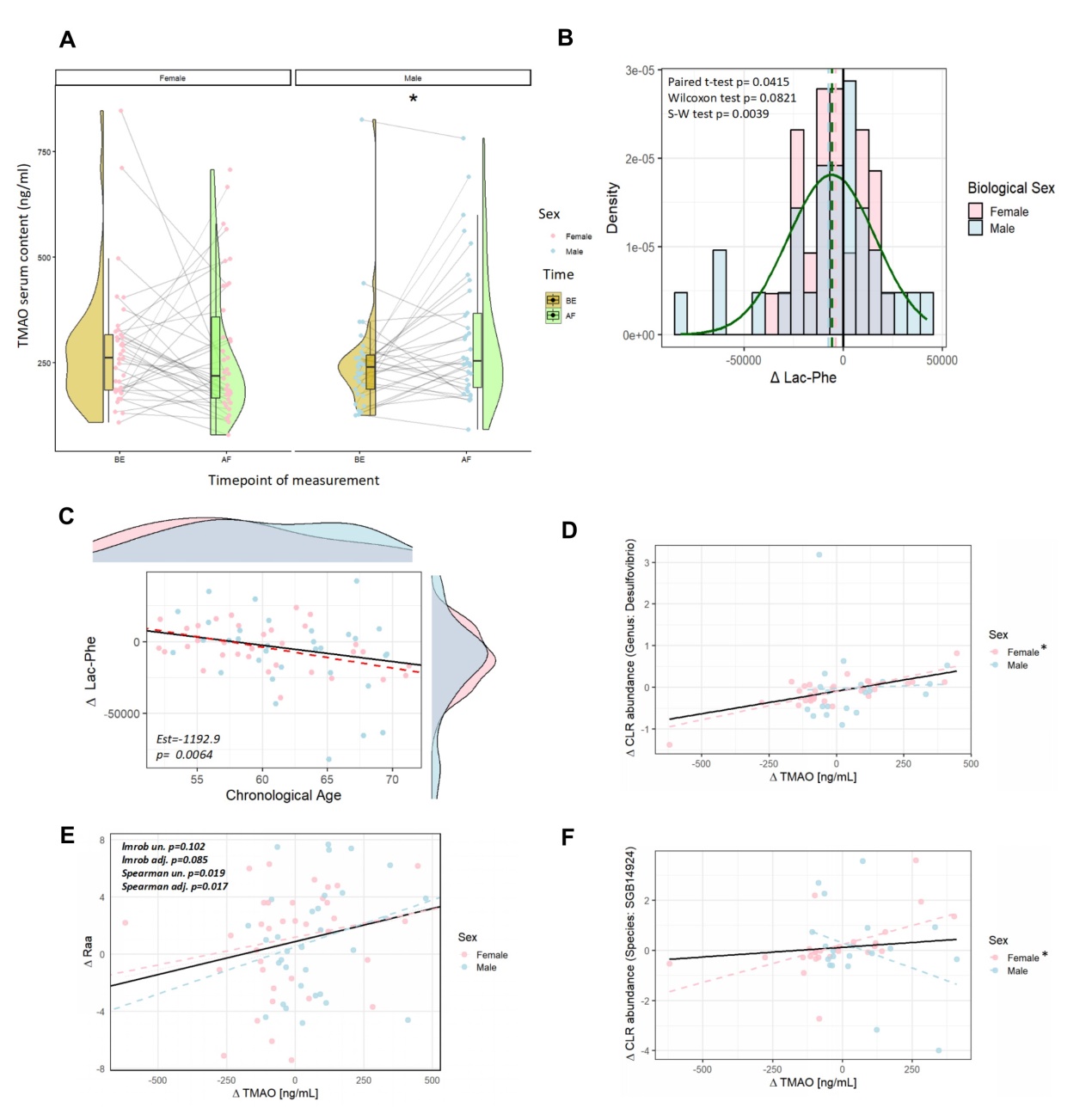

**Supplementary Figure 4. | Changes in Lac-Phe and TMAO During the Exercise Intervention and their Associations with Epigenetic Aging and Related Biomarkers.**

(A) Plasma trimethylamine N-oxide (TMAO) levels increased significantly only in male participants following the exercise intervention. (B) Serum phenyl-lactylation showed a decreasing tendency after the intervention, as assessed by the Wilcoxon signed-rank test; S–W denotes the Shapiro–Wilk normality test. (C) The change in serum phenyl-lactylation (Lac-Phe) was significantly negatively associated with chronological age at baseline (start of the intervention). (E) The change in plasma TMAO showed a significant Spearman rank correlation with the change in right atrial area (RAA). (D, F) The change in TMAO levels showed significant Spearman rank correlations with the relative abundance of the genus Desulfovibrio and SGB14924 (an unclassified microbial species) in female participants. * depicts siginficant sex specific associations at p<0.05.

**Supplementary Tables**

**Supplementary Table 1. | Association of Exercise Volume with Methylation Based Estimates**

| Association of MET with methylation based estimates |  |  |  |  |  |  |
| --- | --- | --- | --- | --- | --- | --- |
| Protein | ρ | P (Spearman) | β | P (Robust) | Adj β | Adj P |
| TGF.alpha_D | −0.388 | **0.000829** | -1.25E-03 | **0.00028** | -1.25E-03 | **0.00032** |
| GZMA_D | 0.36 | **0.00239** | 4.83E-04 | **0.0014** | 4.93E-04 | **0.00088** |
| EN.RAGE_D | −0.328 | **0.00526** | -1.10E-03 | **0.00293** | -1.24E-03 | **0.00089** |
| NK_D | 0.32 | **0.00744** | 2.10E-04 | **0.0033** | 2.05E-04 | **0.00937** |
| HGF_D | −0.328 | **0.0053** | -6.26E-04 | **0.00565** | -6.75E-04 | **0.0026** |
| NK.EpiDISH_D | 0.30 | **0.0103** | 1.13E-04 | **0.00653** | 1.12E-04 | NA |
| acute.upper.respiratory.infections_D | 0.25 | **0.0384** | 9.08E-04 | **0.00879** | 4.83E-04 | 0.159 |
| DNAm_M_HDL_L_D | 0.28 | **0.0178** | 1.74E-03 | **0.0142** | 9.44E-04 | 0.0757 |
| DNAm_M_HDL_C_D | 0.23 | 0.0579 | 1.60E-03 | **0.0354** | 3.49E-04 | 0.558 |
| Gran_D | −0.240 | **0.044** | -2.94E-04 | **0.0438** | -3.18E-04 | **0.0391** |
| DNAm_ApoA1_D | 0.23 | 0.0568 | 1.87E-03 | **0.0459** | 9.25E-04 | **0.0491** |
| Antihistamines...Ethanolamines_D | −0.234 | **0.0493** | -1.77E-03 | **0.048** | -1.05E-03 | **0.0305** |
| Biguanides_D | 0.04 | 0.722 | 2.11E-03 | 0.0512 | 2.07E-03 | **0.0403** |
| Opioid.Antagonists_D | −0.229 | 0.0547 | -2.71E-03 | 0.0553 | -1.23E-03 | **0.0452** |
| OSM_D | −0.224 | 0.0607 | -1.05E-03 | 0.057 | -1.11E-03 | **0.0359** |
| ABSOLUTE.MONO.COUNT_D | 0.18 | 0.138 | 2.41E-04 | 0.0608 | 9.14E-05 | 0.286 |
| GLUCOSE_D | 0.21 | 0.0748 | 1.14E-01 | 0.0632 | 2.42E-02 | 0.388 |
| DNAm_L_VLDL_L_D | 0.25 | **0.0342** | 9.75E-04 | 0.0655 | 1.45E-03 | **0.0161** |
| Bcell_D | 0.21 | 0.0807 | 3.77E-05 | 0.0665 | 4.02E-05 | 0.0758 |
| DNAm_LDL_C_D | −0.173 | 0.148 | -1.06E-03 | 0.0752 | -7.34E-04 | 0.255 |
| DNAm_XL_VLDL_L_D | 0.21 | 0.0858 | 9.39E-04 | 0.0768 | 1.67E-03 | **0.00761** |
| ISCHEMIC.HEART.DISEASE_D | 0.03 | 0.797 | 1.55E-03 | 0.0815 | 1.09E-03 | 0.174 |
| DNAm_MetaboHealth_D | −0.212 | 0.0754 | -6.61E-04 | 0.0839 | -5.18E-04 | 0.0775 |
| NMNAT1_D | 0.22 | 0.0651 | 3.35E-04 | 0.0895 | 1.98E-04 | 0.382 |
| heart.valve.disorders_D | 0.05 | 0.684 | 5.99E-04 | 0.0947 | 5.17E-04 | 0.16 |
| DNAm_Omega_6_perc_D | −0.272 | **0.0216** | -1.41E-03 | 0.0948 | -1.41E-03 | 0.118 |
| DNAm_LDL_particle_size_D | −0.251 | **0.0347** | -1.26E-03 | 0.103 | -1.50E-03 | **0.0493** |
| Other.Immune.Disorders_D | −0.060 | 0.618 | -1.02E-03 | 0.104 | -1.28E-03 | **0.0141** |
| Antiarrhythmics.Type.III_D | 0.21 | 0.0849 | 1.95E-03 | 0.108 | 7.03E-04 | 0.111 |
| Cephalosporins...4th.Generation_D | 0.17 | 0.148 | 2.08E-03 | 0.112 | 7.49E-04 | 0.232 |
| Heparins.And.Heparinoid.Like.Agents_D | −0.189 | 0.115 | -5.23E-04 | 0.115 | -4.74E-04 | 0.0663 |
| Specialty.Vitamins.Products_D | 0.09 | 0.447 | 4.83E-03 | 0.116 | -1.59E-04 | 0.903 |
| FERRITIN_D | 0.20 | 0.0928 | 7.67E-01 | 0.118 | 1.62E-01 | 0.411 |
| DNAm_SFA_perc_D | 0.19 | 0.115 | 1.12E-03 | 0.119 | 4.64E-04 | 0.549 |
| ABSOLUTE.LYMPHOCYTE.COUNT_D | 0.21 | 0.0773 | 1.07E-03 | 0.12 | 6.73E-04 | 0.13 |
| NEUTROPHIL.PERCENT..AUTO_D | −0.192 | 0.109 | -9.49E-03 | 0.123 | -6.55E-03 | 0.176 |
| IRON.BINDING.CAPACITY_D | 0.15 | 0.212 | 3.62E-02 | 0.124 | 8.80E-03 | 0.678 |
| Mono.EpiDISH_D | 0.20 | 0.0946 | 6.77E-05 | 0.128 | 7.72E-05 | **0.0468** |
| DNAm_PUFA_perc_D | −0.254 | **0.0329** | -1.10E-03 | 0.128 | -1.06E-03 | 0.238 |
| Antitussives_D | 0.14 | 0.232 | 2.21E-03 | 0.134 | 7.48E-04 | 0.566 |
| Local.Anesthetic.Combinations_D | −0.175 | 0.144 | -1.87E-03 | 0.135 | -5.42E-04 | 0.238 |
| Phosphate.Binder.Agents_D | −0.161 | 0.181 | -3.25E-03 | 0.141 | -1.08E-03 | 0.433 |
| DNAm_XL_HDL_C_D | −0.219 | 0.0665 | -1.44E-03 | 0.142 | -1.81E-03 | 0.0711 |
| CMV.Agents_D | 0.11 | 0.351 | 3.02E-03 | 0.143 | 7.27E-04 | 0.585 |
| DNAm_Phosphoglycerides_D | 0.22 | 0.0643 | 7.83E-04 | 0.143 | 9.48E-04 | 0.116 |
| Potassium_D | −0.157 | 0.192 | -1.58E-03 | 0.144 | -6.34E-04 | 0.269 |
| Other.diseases.of.digestive.system_D | −0.049 | 0.682 | 1.67E-03 | 0.151 | 3.18E-04 | 0.697 |
| FcRL2_D | 0.13 | 0.27 | 2.81E-04 | 0.156 | 1.97E-04 | 0.222 |
| NEUTROPHILS.ABS..PRELIM._D | −0.173 | 0.149 | -9.58E-04 | 0.16 | -1.43E-03 | **0.0234** |
| GDF.8_D | 0.13 | 0.28 | 1.84E-04 | 0.168 | 1.81E-04 | 0.247 |
| DNAm_MUFA_perc_D | 0.17 | 0.169 | 7.93E-04 | 0.169 | 8.59E-04 | 0.152 |
| Calcium.Channel.Blockers_D | 0.14 | 0.259 | 1.27E-03 | 0.177 | 8.31E-04 | 0.29 |
| DNAm_Albumin_D | 0.15 | 0.215 | 1.04E-03 | 0.178 | 1.25E-03 | 0.142 |
| Sleep.disorders_D | 0.12 | 0.309 | 1.21E-03 | 0.181 | 2.90E-04 | 0.543 |
| Analgesics.Other_D | −0.180 | 0.134 | -2.26E-03 | 0.187 | -7.37E-04 | 0.0692 |
| TOTAL.PROTEIN_D | 0.17 | 0.165 | 2.08E-04 | 0.187 | 1.57E-04 | 0.297 |
| MEAN.CORPUSCULAR.HEMOGLOBIN_D | −0.015 | 0.903 | 1.13E-03 | 0.189 | 1.06E-03 | 0.126 |
| CRTAM_D | 0.14 | 0.235 | 3.65E-04 | 0.192 | 4.05E-04 | 0.119 |
| DNAm_Total_cholines_D | 0.17 | 0.162 | 6.88E-04 | 0.192 | 2.41E-04 | NA |
| Liquid.Vehicles_D | 0.12 | 0.335 | 7.59E-04 | 0.196 | -1.06E-04 | 0.83 |
| DNAm_HDL_C_D | 0.12 | 0.308 | 6.85E-04 | 0.196 | 3.54E-04 | 0.493 |
| HEMATOCRIT_D | −0.161 | 0.181 | -3.00E-03 | 0.198 | -2.45E-03 | 0.299 |
| CREATININE_D | −0.123 | 0.307 | -1.87E-03 | 0.198 | -7.16E-05 | 0.949 |
| Heart.failure_D | −0.028 | 0.817 | -4.86E-04 | 0.199 | -8.50E-04 | 0.088 |
| diseases.of.kidney.and.ureters_D | 0.25 | **0.038** | 3.42E-03 | 0.199 | 1.24E-03 | **0.0423** |
| DNAm_Lactate_D | 0.13 | 0.267 | 7.37E-04 | 0.217 | 5.12E-04 | NA |
| DORSOPATHIES_D | 0.19 | 0.109 | 8.64E-04 | 0.22 | 6.77E-04 | 0.195 |
| Impotence.Agents_D | −0.006 | 0.96 | 1.92E-03 | 0.22 | 1.46E-03 | 0.355 |
| Parenteral.Therapy.Supplies_D | 0.18 | 0.139 | 2.32E-03 | 0.223 | 1.07E-03 | 0.557 |
| Liver.disease_D | 0.12 | 0.301 | 8.37E-04 | 0.224 | 2.86E-04 | 0.582 |
| DNAm_ApoB_D | −0.106 | 0.38 | -1.06E-03 | 0.235 | -5.96E-04 | 0.424 |
| MMP.1_D | −0.127 | 0.292 | -1.66E-04 | 0.235 | -8.86E-05 | 0.552 |
| DNAm_Unsaturation_D | −0.132 | 0.274 | -1.63E-03 | 0.235 | -1.06E-03 | 0.33 |
| DNAm_L_HDL_C_D | 0.12 | 0.308 | 1.46E-03 | 0.237 | -4.36E-05 | 0.953 |
| Cephalosporins...3rd.Generation_D | −0.039 | 0.744 | 6.82E-04 | 0.237 | 1.00E-05 | 0.986 |
| Vasodilators_D | 0.14 | 0.253 | 2.26E-03 | 0.239 | -8.05E-05 | 0.899 |
| DNAm_Phenylalanine_D | 0.13 | 0.289 | 4.89E-04 | 0.24 | 7.36E-04 | 0.0666 |
| Beta.Blockers.Cardio.Selective_D | −0.186 | 0.121 | -1.30E-03 | 0.244 | -1.40E-03 | 0.118 |
| Antiseptics...Mouth.Throat_D | 0.07 | 0.593 | 1.13E-03 | 0.247 | 3.40E-04 | 0.598 |
| Other.diseases.of.the.blood_D | 0.13 | 0.279 | 3.25E-03 | 0.253 | 3.22E-04 | 0.561 |
| DNAm_M_LDL_L_D | −0.107 | 0.376 | -9.64E-04 | 0.254 | -4.43E-05 | 0.959 |
| Diabetic.Supplies_D | 0.15 | 0.227 | 2.55E-03 | 0.256 | 2.60E-04 | 0.749 |
| ABSOLUTE.NEUT.COUNT_D | −0.157 | 0.191 | -1.12E-03 | 0.259 | -2.21E-03 | **0.013** |
| DNAm_Leucine_D | 0.16 | 0.191 | 7.29E-04 | 0.26 | 6.56E-04 | 0.293 |
| X.Other.symptoms..reflex_D | 0.12 | 0.33 | 1.13E-03 | 0.265 | -1.15E-04 | 0.825 |
| Radiographic.Contrast.Media_D | 0.12 | 0.328 | 5.84E-04 | 0.268 | 7.01E-04 | 0.196 |
| DNAm_XS_VLDL_L_D | −0.087 | 0.468 | -8.42E-04 | 0.268 | -5.04E-04 | 0.488 |
| WHITE.BLOOD.CELL.COUNT_D | −0.001 | 0.996 | -1.59E-03 | 0.273 | -1.54E-03 | 0.0611 |
| Nitrates_D | −0.211 | 0.0769 | -1.08E-03 | 0.282 | -1.71E-03 | **0.0293** |
| DNAm_VLDL_particle_size_D | −0.080 | 0.505 | -6.88E-04 | 0.285 | -1.16E-04 | 0.865 |
| Vasopressors_D | −0.163 | 0.173 | -6.80E-04 | 0.286 | -6.95E-04 | 0.103 |
| Bone.Density.Regulators_D | 0.10 | 0.388 | 3.49E-03 | 0.291 | 1.74E-05 | 0.988 |
| Dibenzapines_D | −0.073 | 0.548 | -1.97E-03 | 0.297 | 8.84E-05 | 0.925 |
| Sepsis.and.SIRS_D | −0.057 | 0.64 | -4.34E-04 | 0.298 | 2.21E-04 | 0.361 |
| DNAm_Total_C_D | −0.074 | 0.538 | -6.11E-04 | 0.3 | -2.53E-04 | 0.674 |
| MEAN.CORPUSCULAR.VOLUME_D | 0.09 | 0.437 | 2.84E-03 | 0.302 | 1.40E-03 | 0.434 |
| Alkalinizers_D | −0.168 | 0.161 | -7.95E-04 | 0.308 | -8.14E-04 | 0.252 |
| Electrolyte.Mixtures_D | 0.01 | 0.969 | -6.86E-04 | 0.309 | -7.09E-04 | 0.213 |
| Antacids...Bicarbonate_D | 0.11 | 0.354 | 1.31E-03 | 0.325 | -7.52E-05 | 0.928 |
| ALBUMIN_D | 0.13 | 0.268 | 2.59E-04 | 0.326 | 1.51E-04 | 0.204 |
| Osmotic.Diuretics_D | 0.10 | 0.414 | 1.26E-03 | 0.336 | 4.50E-04 | 0.569 |
| MAGNESIUM_D | −0.090 | 0.457 | -4.61E-05 | 0.336 | -7.03E-06 | 0.84 |
| DNAm_PUFA_D | −0.073 | 0.543 | -7.37E-04 | 0.337 | -5.97E-04 | 0.435 |
| Phosphate_D | 0.12 | 0.338 | 9.74E-04 | 0.339 | 3.55E-04 | 0.44 |
| ALANINE.AMINOTRANSFERASE_D | 0.04 | 0.714 | 2.62E-02 | 0.344 | -6.37E-03 | 0.499 |
| UREA.NITROGEN_D | −0.124 | 0.303 | -1.46E-02 | 0.345 | -1.12E-02 | 0.0967 |
| Antiflatulents_D | 0.02 | 0.863 | -7.78E-04 | 0.345 | -8.76E-04 | 0.172 |
| HMG.CoA.Reductase.Inhibitors_D | −0.114 | 0.343 | -5.54E-04 | 0.351 | -9.43E-04 | 0.121 |
| HGB.A1C...HPLC_D | 0.15 | 0.209 | 4.67E-04 | 0.355 | 4.35E-04 | 0.325 |
| DNAm_Glucose_D | 0.14 | 0.257 | 5.77E-04 | 0.365 | 5.46E-04 | 0.434 |
| Laxative.Combinations_D | −0.113 | 0.35 | -1.12E-03 | 0.369 | -3.80E-04 | 0.561 |
| Salicylates_D | −0.132 | 0.272 | -1.58E-03 | 0.369 | -1.05E-03 | 0.195 |
| DNAm_3_Hydroxybutyrate_D | 0.07 | 0.55 | 5.80E-04 | 0.37 | 3.26E-05 | 0.957 |
| DNAm_SFA_D | 0.19 | 0.11 | 9.48E-04 | 0.385 | 1.40E-03 | 0.154 |
| DNAm_HDL_particle_size_D | 0.08 | 0.49 | 7.80E-04 | 0.385 | 5.66E-04 | 0.524 |
| SODIUM_D | 0.05 | 0.705 | 1.03E-03 | 0.39 | 1.88E-04 | 0.813 |
| Glucocorticosteroids_D | 0.01 | 0.966 | -7.04E-04 | 0.401 | -4.26E-04 | 0.506 |
| CD6_D | 0.10 | 0.403 | 2.12E-04 | 0.403 | 3.16E-04 | 0.244 |
| Plasma.Proteins_D | −0.141 | 0.241 | -6.90E-04 | 0.406 | -5.66E-04 | 0.436 |
| dementia.and.related.cognitive.disorders.symptoms_D | 0.12 | 0.302 | 7.78E-04 | 0.416 | 3.30E-04 | 0.751 |
| Fluoroquinolones_D | 0.06 | 0.638 | 1.12E-03 | 0.426 | -8.39E-05 | 0.858 |
| DNAm_Tyrosine_D | 0.09 | 0.461 | 3.71E-04 | 0.429 | 5.01E-04 | 0.179 |
| Angiotensin.II.Receptor.Antagonists_D | −0.004 | 0.972 | -1.92E-04 | 0.431 | -1.54E-04 | 0.538 |
| Benzodiazepines_D | 0.09 | 0.471 | 8.48E-04 | 0.443 | 6.11E-04 | 0.23 |
| Thiazides.and.Thiazide.Like.Diuretics_D | 0.11 | 0.377 | 9.76E-04 | 0.443 | -2.42E-04 | 0.613 |
| CD4T_D | 0.11 | 0.355 | 5.72E-05 | 0.45 | 6.69E-05 | 0.408 |
| Obesity_D | 0.12 | 0.315 | 2.95E-03 | 0.453 | -6.04E-04 | 0.278 |
| Metabolic.Modifiers_D | −0.081 | 0.5 | -1.61E-03 | 0.467 | 1.75E-05 | 0.977 |
| PlasmaBlast_D | −0.061 | 0.615 | -2.71E-04 | 0.468 | 8.33E-06 | 0.973 |
| noninfective.gastrointestinal.disorders_D | 0.14 | 0.26 | 5.25E-04 | 0.469 | 3.43E-04 | 0.605 |
| DNAm_LA_D | −0.049 | 0.683 | -4.24E-04 | 0.471 | 1.43E-05 | 0.983 |
| EZR_D | 0.10 | 0.407 | 1.34E-04 | 0.487 | 1.46E-04 | 0.388 |
| DNAm_DHA_D | 0.10 | 0.403 | 4.56E-04 | 0.496 | 5.53E-05 | 0.922 |
| Antiperistaltic.Agents_D | −0.114 | 0.346 | -1.52E-03 | 0.496 | -6.73E-04 | 0.115 |
| Sympathomimetics_D | −0.066 | 0.584 | -9.28E-04 | 0.497 | 6.09E-04 | 0.353 |
| CXCL9_D | −0.133 | 0.269 | -1.40E-04 | 0.497 | -1.05E-04 | 0.662 |
| Potassium.Removing.Agents_D | −0.143 | 0.235 | -1.09E-03 | 0.5 | -1.23E-03 | 0.139 |
| DNAm_S_HDL_C_D | 0.13 | 0.283 | 7.25E-04 | 0.505 | 5.91E-04 | 0.58 |
| DNAm_Alanine_D | 0.07 | 0.545 | 3.99E-04 | 0.507 | 6.57E-04 | 0.247 |
| Mono_D | −0.015 | 0.903 | 2.24E-05 | 0.51 | 1.41E-05 | 0.676 |
| Anti.infective.Agents...Misc._D | −0.023 | 0.848 | -1.15E-03 | 0.511 | 5.35E-04 | 0.252 |
| Immunosuppressive.Agents_D | 0.04 | 0.765 | -5.67E-04 | 0.515 | -1.18E-03 | 0.215 |
| ABSOLUTE.BASO.COUNT_D | 0.20 | 0.0871 | 3.70E-05 | 0.521 | 1.01E-05 | 0.233 |
| DNAm_Omega_3_D | 0.09 | 0.479 | 2.71E-04 | 0.523 | 6.60E-04 | 0.108 |
| DISEASES.OF.ARTERIES..ARTERIOLES..AND.CAPILLARIES_D | 0.08 | 0.523 | -1.79E-04 | 0.531 | -6.64E-06 | 0.981 |
| HYPERTENSIVE.DISEASE_D | 0.14 | 0.263 | 3.89E-04 | 0.534 | 6.99E-04 | 0.0909 |
| DNAm_Total_triglycerides_D | 0.08 | 0.505 | 4.32E-04 | 0.538 | 9.37E-04 | 0.0649 |
| DNAm_Omega_6_D | −0.022 | 0.854 | -4.26E-04 | 0.538 | -1.47E-04 | 0.833 |
| Antiarrhythmics.Type.I.B_D | −0.044 | 0.713 | -4.35E-04 | 0.545 | 5.05E-05 | 0.933 |
| Imidazole.Related.Antifungals_D | 0.01 | 0.93 | -6.55E-04 | 0.555 | -1.24E-03 | 0.333 |
| Platelet.Aggregation.Inhibitors_D | −0.074 | 0.539 | -8.36E-04 | 0.561 | -3.94E-04 | 0.477 |
| Disorders.of.mineral.metabolism_D | 0.16 | 0.182 | 2.70E-03 | 0.562 | 7.12E-04 | 0.354 |
| Hematopoietic.Growth.Factors_D | 0.12 | 0.327 | 1.20E-03 | 0.564 | -1.54E-03 | 0.386 |
| Anemias_D | −0.090 | 0.458 | -3.22E-04 | 0.569 | -5.06E-04 | 0.237 |
| DNAm_Creatinine_D | 0.08 | 0.488 | 7.28E-04 | 0.572 | 9.50E-04 | 0.328 |
| Anti.infective.Misc....Combinations_D | 0.02 | 0.845 | 8.00E-04 | 0.574 | -8.49E-05 | 0.915 |
| DNAm_IDL_L_D | −0.068 | 0.574 | -5.83E-04 | 0.574 | -4.35E-04 | 0.409 |
| VENTRICULAR.RATE_D | 0.00 | 0.989 | -2.13E-03 | 0.575 | -6.12E-03 | **0.0385** |
| X.chronic.obstructive.pulmonary.disease..bronchiectasis..asthma_D | 0.09 | 0.47 | 4.29E-04 | 0.576 | -1.66E-04 | 0.793 |
| Gallstone.Solubilizing.Agents_D | 0.09 | 0.467 | 1.46E-03 | 0.577 | -5.70E-04 | 0.735 |
| Disorders.of.fluid..electrolyte..and.acid.base.balance_D | −0.018 | 0.878 | 9.82E-04 | 0.577 | -3.34E-04 | 0.355 |
| Phenothiazines_D | 0.05 | 0.656 | 2.08E-03 | 0.578 | -4.32E-04 | 0.703 |
| Malignant.neoplasm.of.skin_D | 0.05 | 0.697 | 6.66E-04 | 0.581 | -2.10E-04 | 0.205 |
| Alternative.Medicine...M.s_D | 0.11 | 0.35 | 2.41E-04 | 0.582 | 2.25E-04 | 0.622 |
| Surfactant.Laxatives_D | 0.01 | 0.917 | -4.01E-04 | 0.585 | -5.36E-04 | 0.237 |
| Posterior.Pituitary.Hormones_D | 0.06 | 0.611 | 1.76E-04 | 0.585 | 2.09E-04 | 0.462 |
| Rheumatism..Excluding.The.Back_D | 0.06 | 0.627 | -5.28E-04 | 0.591 | 7.71E-05 | 0.949 |
| DNAm_S_HDL_L_D | 0.11 | 0.346 | 4.91E-04 | 0.592 | -3.45E-04 | 0.72 |
| DNAm_MUFA_D | 0.13 | 0.269 | 2.74E-04 | 0.604 | 8.49E-04 | 0.123 |
| CD4CD8Ratio_D | 0.13 | 0.329 | 1.55E-03 | 0.607 | 1.14E-03 | 0.677 |
| Other.peri...endo...and.myocarditis_D | −0.010 | 0.934 | 6.28E-04 | 0.614 | -1.27E-03 | 0.0563 |
| ABSOLUTE.EOS.COUNT_D | 0.13 | 0.294 | 2.07E-04 | 0.624 | 8.64E-05 | 0.499 |
| Penicillin.Combinations_D | −0.129 | 0.283 | -7.55E-04 | 0.625 | 3.83E-05 | 0.959 |
| DNAm_XL_HDL_L_D | −0.097 | 0.423 | -4.33E-04 | 0.626 | -1.40E-03 | 0.14 |
| Other.symptoms.of.respiratory.system_D | 0.00 | 0.999 | 1.03E-03 | 0.627 | -5.49E-04 | 0.34 |
| Thrombolytic.Enzymes_D | −0.031 | 0.799 | -6.16E-04 | 0.627 | 3.55E-04 | 0.639 |
| DNAm_L_LDL_L_D | −0.098 | 0.418 | -2.35E-04 | 0.646 | -3.05E-04 | 0.613 |
| DNAm_Acetate_D | 0.04 | 0.72 | 3.96E-04 | 0.647 | 1.68E-04 | 0.789 |
| CD4.naive_D | −0.038 | 0.752 | -2.17E-01 | 0.651 | 1.64E-01 | 0.076 |
| DNAm_XXL_VLDL_L_D | −0.014 | 0.908 | -4.38E-04 | 0.654 | 8.66E-04 | 0.245 |
| X.congenital.anomalies..circulatory.and.cardiac_D | 0.01 | 0.969 | -1.16E-04 | 0.657 | -2.31E-04 | 0.417 |
| Proton.Pump.Inhibitors_D | 0.08 | 0.492 | 3.25E-04 | 0.66 | 3.63E-04 | 0.631 |
| PFT.FEV_1.PRE_D | 0.01 | 0.937 | -1.73E-04 | 0.668 | 2.89E-04 | 0.213 |
| Diagnostic.Radiopharmaceuticals_D | 0.03 | 0.814 | 2.84E-04 | 0.669 | -5.02E-04 | 0.448 |
| Glycopeptides_D | 0.02 | 0.879 | 1.67E-04 | 0.673 | 6.32E-06 | 0.987 |
| CD8T_D | 0.04 | 0.719 | 1.38E-05 | 0.676 | 2.23E-05 | 0.526 |
| ANION.GAP_D | 0.02 | 0.89 | 6.29E-04 | 0.679 | -1.12E-03 | 0.0698 |
| HEMATOCRIT..OSL._D | −0.007 | 0.954 | 3.39E-04 | 0.682 | 1.53E-03 | 0.189 |
| DNAm_Valine_D | 0.01 | 0.933 | -2.93E-04 | 0.685 | -1.00E-05 | 0.988 |
| COAGULATION.OR.HEMORRHAGIC.DISEASES_D | 0.13 | 0.297 | 1.98E-03 | 0.685 | 4.78E-04 | 0.675 |
| Stimulant.Laxatives_D | 0.16 | 0.183 | 1.42E-03 | 0.685 | 5.93E-04 | 0.229 |
| Magnesium_D | −0.144 | 0.23 | -4.98E-04 | 0.69 | -1.32E-04 | 0.807 |
| Cobalamins_D | 0.08 | 0.534 | 5.17E-04 | 0.69 | -4.89E-04 | 0.325 |
| Cataract_D | 0.18 | 0.132 | 5.31E-04 | 0.697 | 3.56E-04 | 0.291 |
| Bacterial.Vaccines_D | 0.16 | 0.177 | 3.12E-04 | 0.7 | 8.52E-04 | 0.145 |
| SIGLEC1_D | −0.105 | 0.386 | -8.75E-05 | 0.704 | -2.22E-04 | 0.254 |
| CHOLESTEROL..HDL_D | 0.03 | 0.797 | -1.20E-03 | 0.705 | -2.26E-03 | 0.462 |
| cardiac.conduction.disorders_D | −0.097 | 0.42 | 1.20E-04 | 0.708 | 1.76E-04 | NA |
| Diagnostic.Drugs_D | −0.056 | 0.643 | -7.18E-04 | 0.71 | -4.10E-04 | 0.457 |
| DNAm_L_HDL_L_D | 0.03 | 0.826 | 2.83E-04 | 0.72 | -9.08E-04 | 0.196 |
| DNAm_Total_fatty_acids_D | 0.11 | 0.345 | 3.64E-04 | 0.722 | 8.49E-04 | 0.411 |
| Saline.Laxatives_D | 0.04 | 0.733 | 2.97E-04 | 0.731 | 2.77E-04 | 0.702 |
| DNAm_Glutamine_D | −0.087 | 0.47 | -2.82E-04 | 0.733 | -1.04E-04 | 0.886 |
| HEMOGLOBIN_D | −0.036 | 0.763 | 3.54E-04 | 0.734 | 5.46E-04 | 0.634 |
| DNAm_Histidine_D | 0.09 | 0.472 | 1.85E-04 | 0.738 | 1.73E-04 | 0.782 |
| Gout.Agents_D | 0.03 | 0.777 | 2.96E-04 | 0.747 | -4.53E-05 | 0.959 |
| B.Complex.w..Folic.Acid_D | 0.04 | 0.728 | 2.38E-04 | 0.747 | 2.12E-04 | 0.804 |
| DNAm_Isoleucine_D | 0.03 | 0.793 | 1.53E-04 | 0.749 | 3.69E-04 | 0.516 |
| SKR3_D | −0.086 | 0.474 | -1.08E-04 | 0.75 | -2.42E-04 | 0.534 |
| Prostatic.Hypertrophy.Agents_D | 0.05 | 0.695 | 2.15E-04 | 0.754 | -2.48E-04 | 0.718 |
| osteoarthritis_D | 0.07 | 0.566 | 2.59E-04 | 0.759 | -4.03E-04 | 0.6 |
| DNAm_Sphingomyelins_D | 0.00 | 0.973 | -2.20E-04 | 0.764 | 1.35E-04 | 0.754 |
| DNAm_M_VLDL_L_D | 0.04 | 0.768 | -2.05E-04 | 0.77 | 5.25E-04 | 0.4 |
| DNAm_S_LDL_L_D | 0.03 | 0.828 | -1.94E-04 | 0.782 | 5.12E-04 | 0.453 |
| Thyroid.Hormones_D | 0.09 | 0.452 | 1.35E-04 | 0.788 | 1.25E-04 | 0.803 |
| Anti.infectives...Throat_D | −0.132 | 0.273 | -1.18E-03 | 0.796 | -8.85E-04 | 0.211 |
| DNAm_IDL_C_D | −0.020 | 0.87 | -1.72E-04 | 0.819 | -1.61E-04 | 0.784 |
| NEP_D | 0.02 | 0.855 | 6.11E-05 | 0.827 | -1.21E-04 | 0.671 |
| Poisoning.By.Drugs..Medicinal.And.Biological.Substances_D | 0.03 | 0.796 | 1.93E-04 | 0.827 | -3.54E-05 | 0.964 |
| CHLORIDE_D | 0.10 | 0.432 | 1.79E-03 | 0.828 | 8.36E-05 | 0.933 |
| PLATELET.COUNT..AUTO_D | 0.08 | 0.495 | -8.47E-03 | 0.83 | -2.65E-02 | 0.391 |
| Antacid.Combinations_D | 0.04 | 0.746 | -1.34E-04 | 0.833 | -5.07E-04 | 0.319 |
| DIABETES_D | 0.07 | 0.576 | 1.33E-04 | 0.834 | 1.41E-04 | 0.822 |
| SMPD1_D | 0.00 | 0.997 | 7.64E-05 | 0.838 | -1.81E-04 | 0.683 |
| ALKALINE.PHOSPHATASE_D | 0.03 | 0.788 | 1.31E-02 | 0.848 | -5.22E-03 | 0.521 |
| CD8pCD28nCD45RAn_D | −0.012 | 0.92 | -1.22E-03 | 0.85 | -7.77E-03 | 0.261 |
| ABSOLUTE.IMMATURE.GRAN.COUNT_D | 0.06 | 0.605 | 5.97E-06 | 0.852 | -1.34E-07 | 0.996 |
| Alpha.Beta.Blockers_D | −0.084 | 0.488 | 1.61E-04 | 0.865 | -5.87E-05 | 0.934 |
| DNAm_S_VLDL_L_D | 0.08 | 0.491 | 1.51E-04 | 0.867 | 3.60E-04 | 0.705 |
| DNAm_Acetoacetate_D | −0.009 | 0.939 | -1.38E-04 | 0.869 | 4.91E-04 | 0.512 |
| QRS.DURATION_D | 0.04 | 0.769 | -9.43E-04 | 0.877 | -4.88E-03 | 0.409 |
| DNAm_Citrate_D | 0.02 | 0.866 | 1.74E-04 | 0.892 | 1.02E-03 | 0.252 |
| Iron_D | −0.010 | 0.937 | 6.60E-04 | 0.892 | -1.54E-03 | 0.123 |
| Insulin_D | −0.073 | 0.547 | 8.97E-05 | 0.893 | 4.19E-05 | 0.925 |
| Disorders.of.lipoid.metabolism_D | −0.148 | 0.218 | -5.86E-05 | 0.897 | -6.05E-04 | 0.087 |
| X.osteopenia..osteoporosis..pathological.fractures_D | 0.06 | 0.633 | -9.55E-05 | 0.897 | 1.20E-03 | NA |
| DNAm_Glycoprotein_acetyls_D | 0.03 | 0.819 | 9.00E-05 | 0.899 | 3.12E-04 | 0.673 |
| PAIN_D | 0.05 | 0.693 | 6.78E-05 | 0.916 | -1.66E-04 | 0.801 |
| DNAm_Omega_3_perc_D | 0.02 | 0.862 | 4.40E-05 | 0.921 | -9.70E-05 | 0.824 |
| CHOLESTEROL_D | 0.09 | 0.473 | 1.70E-03 | 0.922 | 7.85E-03 | 0.62 |
| Antibiotics...Topical_D | 0.08 | 0.528 | 9.15E-05 | 0.931 | 5.64E-05 | 0.894 |
| Carbohydrates_D | −0.023 | 0.852 | -6.83E-05 | 0.931 | -1.36E-04 | 0.798 |
| FGF.21_D | −0.007 | 0.955 | 2.76E-05 | 0.951 | -1.60E-04 | 0.724 |
| CD8.naive_D | 0.01 | 0.969 | -2.56E-03 | 0.951 | -4.74E-02 | 0.357 |
| DNAm_Phosphatidylcholines_D | 0.06 | 0.605 | 3.87E-05 | 0.951 | 2.27E-04 | 0.716 |
| Pneumoconioses.and.other.lung.diseases_D | 0.02 | 0.842 | 8.39E-05 | 0.956 | 2.88E-04 | 0.714 |
| Arthropathies_D | 0.09 | 0.437 | -3.95E-05 | 0.957 | -9.74E-05 | 0.889 |
| X5.HT3.Receptor.Antagonists_D | 0.01 | 0.926 | 4.46E-04 | 0.96 | -1.17E-03 | 0.134 |
| Loop.Diuretics_D | −0.099 | 0.412 | -2.30E-05 | 0.971 | -3.32E-04 | 0.654 |
| DNAm_VLDL_C_D | 0.05 | 0.701 | -3.51E-05 | 0.974 | 4.78E-04 | 0.533 |
| Laxatives...Miscellaneous_D | 0.06 | 0.62 | -1.31E-05 | 0.983 | 5.39E-04 | NA |
| SEDIMENTATION.RATE..ERYTHROCYTE_D | 0.05 | 0.693 | 2.29E-04 | 0.984 | -5.96E-03 | 0.561 |
| Calcium_D | 0.06 | 0.607 | 8.14E-06 | 0.99 | 2.86E-04 | 0.611 |
| BCAM_D | 0.10 | 0.412 | 0.00E+00 | NA | NA | NA |
| MMP.2_D | −0.099 | 0.412 | 0.00E+00 | NA | NA | NA |
| Lysozyme_D | −0.099 | 0.412 | 0.00E+00 | NA | NA | NA |

The table depicts the associations between cumulative MET-hours and DNA methylation-based estimates. The first column indicates the pre- to post-intervention variables. ρ and P (Spearman) represent the Spearman correlation coefficient and its corresponding p-value for the nonparametric association. β and P (Robust) denote the robust regression coefficient and its corresponding p-value. Adj. β represents the robust regression coefficient adjusted for biological sex, chronological age, and batch effects, together with its corresponding p-value. Red numbers indicate statistically significant associations.

**Supplementary Table 2. | Association of Exercise Volume with Robust GrimAge Realted Metylation Surrogate Biomarkers**

| Association Results With Total MET-hrs |  |  |  |  |  |  |
| --- | --- | --- | --- | --- | --- | --- |
| Spearman and Robust Regression Analyses |  |  |  |  |  |  |
| Variable | Spearman ρ | p (Spearman) | β | p (Robust) | Adj β | Adj p |
| RobustTIMP1_diff | −0.246 | **3.85 × 10^−2^** | −0.584 | **4.61 × 10^−2^** | −0.728 | **1.61 × 10^−2^** |
| RobustPAI1_diff | −0.191 | 1.11 × 10^−1^ | −1.528 | 6.34 × 10^−2^ | −1.541 | 5.46 × 10^−2^ |
| RobustB2M_diff | −0.155 | 1.98 × 10^−1^ | −39.226 | 2.27 × 10^−1^ | −52.313 | 9.22 × 10^−2^ |
| RobustADM_diff | −0.243 | **4.09 × 10^−2^** | −0.019 | **1.09 × 10^−2^** | −0.010 | 1.07 × 10^−1^ |
| RobustCystatinC_diff | −0.078 | 5.17 × 10^−1^ | −4.934 | 5.98 × 10^−1^ | −15.720 | 1.09 × 10^−1^ |
| RobustPACKYRS_diff | −0.166 | 1.65 × 10^−1^ | −0.005 | 8.08 × 10^−2^ | −0.005 | 1.69 × 10^−1^ |
| RobustGDF15_diff | −0.046 | 7.05 × 10^−1^ | −0.012 | 7.35 × 10^−1^ | −0.048 | 2.04 × 10^−1^ |
| RobustLeptin_diff | 0.077 | 5.25 × 10^−1^ | 0.305 | 4.13 × 10^−1^ | 0.402 | 2.45 × 10^−1^ |
| Association Results With Robust DNAm ADM |  |  |  |  |  |  |
| Spearman and Robust Regression Analyses |  |  |  |  |  |  |
| Variable | Spearman ρ | p (Spearman) | β | p (Robust) | Adj β | Adj p |
| Diff_MQ_card | 0.205 | 8.93 × 10^−2^ | 0.245 | **4.94 × 10^−2^** | 0.493 | **2.79 × 10^−2^** |
| Diff_PULZ_card | 0.099 | 4.16 × 10^−1^ | 0.327 | 3.00 × 10^−1^ | 0.764 | **3.23 × 10^−2^** |
| Diff_Rel.LVWT_card | 0.232 | 5.34 × 10^−2^ | 0.061 | 1.05 × 10^−1^ | 0.098 | 6.63 × 10^−2^ |
| Diff_latE..A._card | −0.211 | 7.97 × 10^−2^ | −0.023 | **4.74 × 10^−2^** | −0.020 | 8.34 × 10^−2^ |
| Diff_GOT_blood | 0.106 | 3.78 × 10^−1^ | 0.089 | 4.10 × 10^−1^ | 0.249 | 1.64 × 10^−1^ |
| Diff_GPT_blood | 0.114 | 3.45 × 10^−1^ | 0.249 | 2.72 × 10^−1^ | 0.428 | 2.11 × 10^−1^ |
| Diff_TAPSE_card | 0.088 | 5.02 × 10^−1^ | 0.11 | 5.20 × 10^−1^ | −0.263 | 2.42 × 10^−1^ |
| Diff_VCF_card | −0.071 | 5.60 × 10^−1^ | −0.006 | 3.40 × 10^−1^ | −0.011 | 2.68 × 10^−1^ |
| Diff_latS._card | 0.135 | 2.72 × 10^−1^ | 0.001 | 1.30 × 10^−1^ | 0.001 | 2.76 × 10^−1^ |
| Diff_medE..A._card | −0.101 | 4.05 × 10^−1^ | −0.011 | 3.51 × 10^−1^ | −0.013 | 3.30 × 10^−1^ |
| Diff_BPDIAST_card | −0.076 | 5.34 × 10^−1^ | −0.008 | 9.78 × 10^−1^ | 0.303 | 3.39 × 10^−1^ |
| Diff_Rel.LVID_card | 0.07 | 5.65 × 10^−1^ | −0.012 | 9.20 × 10^−1^ | −0.164 | 3.53 × 10^−1^ |
| Diff_Relsyst_card | 0.104 | 3.92 × 10^−1^ | 0.201 | 2.38 × 10^−1^ | 0.187 | 3.59 × 10^−1^ |
| Diff_GGT_blood | 0.062 | 6.09 × 10^−1^ | 0.133 | 3.63 × 10^−1^ | 0.195 | 4.18 × 10^−1^ |
| Diff_HDL_blood | 0.203 | 8.88 × 10^−2^ | 0.012 | **1.54 × 10^−2^** | 0.007 | 4.65 × 10^−1^ |
| Diff_medS._card | −0.080 | 5.13 × 10^−1^ | 0 | 9.83 × 10^−1^ | 0 | 5.76 × 10^−1^ |
| Diff_BPSYST_card | −0.156 | 1.98 × 10^−1^ | −0.550 | 2.68 × 10^−1^ | −0.460 | 6.19 × 10^−1^ |
| Diff_LDL_blood | −0.508 | **6.18 × 10^−6^** | −0.103 | **5.41 × 10^−6^** | 0.011 | 6.56 × 10^−1^ |
| Diff_Tg_blood | −0.063 | 6.02 × 10^−1^ | 0 | 9.85 × 10^−1^ | 0.004 | 6.76 × 10^−1^ |
| Diff_LVMM_card | 0.101 | 4.07 × 10^−1^ | 0.808 | 7.09 × 10^−1^ | 0.744 | 7.37 × 10^−1^ |
| Diff_RVDA_card | −0.029 | 8.13 × 10^−1^ | −0.146 | 4.61 × 10^−1^ | −0.074 | 7.38 × 10^−1^ |
| Diff_E.A_card | −0.047 | 7.02 × 10^−1^ | −0.001 | 8.27 × 10^−1^ | 0.003 | 7.43 × 10^−1^ |
| Diff_LAa_card | −0.022 | 8.56 × 10^−1^ | −0.017 | 8.63 × 10^−1^ | −0.047 | 7.57 × 10^−1^ |
| Diff_LVET.QT_card | 0.115 | 3.49 × 10^−1^ | 0.002 | 6.68 × 10^−1^ | −0.001 | 7.58 × 10^−1^ |
| Diff_E.medE._card | 0.097 | 4.25 × 10^−1^ | 0.063 | 3.28 × 10^−1^ | −0.026 | 8.26 × 10^−1^ |
| Diff_CON_N_card | 0.093 | 4.42 × 10^−1^ | 0.012 | 5.40 × 10^−1^ | −0.006 | 8.62 × 10^−1^ |
| Diff_Glu_blood | −0.119 | 3.22 × 10^−1^ | −0.004 | 7.67 × 10^−1^ | 0.001 | 9.54 × 10^−1^ |
| Diff_E.latE._card | 0.107 | 3.79 × 10^−1^ | 0.054 | 3.78 × 10^−1^ | −0.004 | 9.56 × 10^−1^ |
| Diff_Chol_blood | 0.035 | 7.73 × 10^−1^ | 0.004 | 8.13 × 10^−1^ | 0.001 | 9.62 × 10^−1^ |
| Diff_Raa_card | 0.125 | 3.01 × 10^−1^ | 0.065 | 5.22 × 10^−1^ | 0.003 | 9.81 × 10^−1^ |
| Diff_LVMM_N_card*^1^* | *1* 0.232 | *1* 5.32 × 10^−2^ | *1* 0.721 | *1* 3.34 × 10^−1^ | *1* 0.502 | *1* * |
| Association Results With Robust DNAm TIMP1 |  |  |  |  |  |  |
| Spearman and Robust Regression Analyses |  |  |  |  |  |  |
| Variable | Spearman ρ | p (Spearman) | β | p (Robust) | Adj β | Adj p |
| Diff_PULZ_card | 0.285 | **1.70 × 10^−2^** | 0.019 | **2.84 × 10^−3^** | 0.019 | **5.79 × 10^−4^** |
| Diff_latE..A._card | −0.304 | **1.05 × 10^−2^** | −0.001 | **6.21 × 10^−3^** | −0.001 | **1.05 × 10^−2^** |
| Diff_GGT_blood | 0.08 | 5.10 × 10^−1^ | 0.007 | **4.32 × 10^−2^** | 0.007 | 6.20 × 10^−2^ |
| Diff_Rel.LVWT_card | 0.272 | **2.27 × 10^−2^** | 0.002 | **4.71 × 10^−2^** | 0.002 | 7.35 × 10^−2^ |
| Diff_MQ_card | 0.331 | **5.11 × 10^−3^** | 0.01 | 7.97 × 10^−2^ | 0.011 | 1.04 × 10^−1^ |
| Diff_BPDIAST_card | −0.024 | 8.46 × 10^−1^ | 0.005 | 4.09 × 10^−1^ | 0.009 | 1.62 × 10^−1^ |
| Diff_GOT_blood | 0.112 | 3.51 × 10^−1^ | 0.004 | 2.80 × 10^−1^ | 0.005 | 1.74 × 10^−1^ |
| Diff_LVMM_card | 0.091 | 4.53 × 10^−1^ | 0.036 | 2.03 × 10^−1^ | 0.042 | 1.88 × 10^−1^ |
| Diff_medS._card | 0.015 | 9.03 × 10^−1^ | 0 | 3.51 × 10^−1^ | 0 | 2.06 × 10^−1^ |
| Diff_medE..A._card | −0.121 | 3.20 × 10^−1^ | 0 | 1.93 × 10^−1^ | 0 | 2.15 × 10^−1^ |
| Diff_LDL_blood | −0.058 | 6.33 × 10^−1^ | 0 | 8.57 × 10^−1^ | 0.001 | 2.61 × 10^−1^ |
| Diff_GPT_blood | 0.127 | 2.90 × 10^−1^ | 0.009 | 1.95 × 10^−1^ | 0.01 | 2.73 × 10^−1^ |
| Diff_latS._card | 0.06 | 6.28 × 10^−1^ | 0 | 3.42 × 10^−1^ | 0 | 3.24 × 10^−1^ |
| Diff_Tg_blood | 0.058 | 6.32 × 10^−1^ | 0 | 3.88 × 10^−1^ | 0 | 3.40 × 10^−1^ |
| Diff_E.A_card | −0.162 | 1.79 × 10^−1^ | 0 | 1.42 × 10^−1^ | 0 | 3.54 × 10^−1^ |
| Diff_HDL_blood | 0.167 | 1.65 × 10^−1^ | 0 | 5.76 × 10^−2^ | 0 | 3.79 × 10^−1^ |
| Diff_Chol_blood | 0.059 | 6.25 × 10^−1^ | 0 | 5.51 × 10^−1^ | 0 | 4.56 × 10^−1^ |
| Diff_Relsyst_card | 0.07 | 5.64 × 10^−1^ | 0.005 | 2.75 × 10^−1^ | 0.003 | 4.75 × 10^−1^ |
| Diff_BPSYST_card | −0.022 | 8.57 × 10^−1^ | 0.007 | 6.83 × 10^−1^ | 0.012 | 4.80 × 10^−1^ |
| Diff_Rel.LVID_card | −0.133 | 2.71 × 10^−1^ | −0.001 | 6.83 × 10^−1^ | −0.002 | 5.38 × 10^−1^ |
| Diff_Raa_card | 0.034 | 7.80 × 10^−1^ | 0.001 | 6.54 × 10^−1^ | 0.001 | 6.16 × 10^−1^ |
| Diff_LVET.QT_card | 0.014 | 9.09 × 10^−1^ | 0 | 9.09 × 10^−1^ | 0 | 6.69 × 10^−1^ |
| Diff_CON_N_card | −0.008 | 9.48 × 10^−1^ | 0 | 5.64 × 10^−1^ | 0 | 7.18 × 10^−1^ |
| Diff_LAa_card | −0.051 | 6.74 × 10^−1^ | 0 | 9.44 × 10^−1^ | 0.001 | 7.61 × 10^−1^ |
| Diff_E.medE._card | −0.071 | 5.60 × 10^−1^ | 0 | 9.50 × 10^−1^ | 0 | 8.31 × 10^−1^ |
| Diff_VCF_card | −0.050 | 6.81 × 10^−1^ | 0 | 8.23 × 10^−1^ | 0 | 9.38 × 10^−1^ |
| Diff_RVDA_card | 0.043 | 7.26 × 10^−1^ | 0 | 9.90 × 10^−1^ | 0 | 9.49 × 10^−1^ |
| Diff_Glu_blood | −0.127 | 2.91 × 10^−1^ | 0 | 9.31 × 10^−1^ | 0 | 9.91 × 10^−1^ |
| Diff_LVMM_N_card*^1^* | *1* 0.118 | *1* 3.31 × 10^−1^ | *1* 0.014 | *1* 1.54 × 10^−1^ | *1* 0.015 | *1* * |
| Diff_E.latE._card*^1^* | *1* 0.071 | *1* 5.60 × 10^−1^ | *1* 0.001 | *1* 6.73 × 10^−1^ | *1* 0.000 | *1* * |
| Diff_TAPSE_card*^1^* | *1* −0.110 | *1* 4.02 × 10^−1^ | *1* −0.002 | *1* 5.66 × 10^−1^ | *1* −0.004 | *1* * |
| *^1^* *: S refinements did not converge |  |  |  |  |  |  |

The table presents the associations between cumulative MET-hours and Robust GrimAge-related DNA methylation estimates, followed by the biomarkers associated with cumulative MET-hours and their relationships with changes in physiological markers. The first column indicates the pre- to post-intervention variables. ρ and P (Spearman) represent the Spearman correlation coefficient and its corresponding p-value for the nonparametric association. β and P (Robust) denote the robust regression coefficient and its corresponding p-value. Adj. β represents the robust regression coefficient adjusted for biological sex, chronological age, and batch effects, together with its corresponding p-value. Red numbers indicate statistically significant associations.

**Supplementary Table 3. | Significant changes in microbial metabolic pathways following the 6-month exercise intervention.**

| Significant Bacterial Pathway Changes (Pre vs Post) Wilcoxon paired comparison in all participants |  |  |  |  |  |
| --- | --- | --- | --- | --- | --- |
| Pathway | Group | N pairs | log2 Fold Change | Wilcoxon p | FDR-adjusted p |
| **UDPNAGSYN-PWY: UDP-N-acetyl-D-glucosamine biosynthesis I** | **All** | **52** | **0.36** | **1.11 × 10^−4^** | **1.60 × 10^−2^** |
| **OANTIGEN-PWY: O-antigen building blocks biosynthesis (E. coli)** | **All** | **52** | **0.27** | **1.60 × 10^−4^** | **1.60 × 10^−2^** |
| **ARGSYNBSUB-PWY: L-arginine biosynthesis II (acetyl cycle)** | **All** | **52** | **0.29** | **1.66 × 10^−4^** | **1.60 × 10^−2^** |
| **COMPLETE-ARO-PWY: superpathway of aromatic amino acid biosynthesis** | **All** | **52** | **0.13** | **1.92 × 10^−4^** | **1.60 × 10^−2^** |
| **GLUTORN-PWY: L-ornithine biosynthesis I** | **All** | **52** | **0.27** | **1.99 × 10^−4^** | **1.60 × 10^−2^** |
| **PWY0-1296: purine ribonucleosides degradation** | **All** | **52** | **0.23** | **2.06 × 10^−4^** | **1.60 × 10^−2^** |
| **ARGSYN-PWY: L-arginine biosynthesis I (via L-ornithine)** | **All** | **52** | **0.26** | **2.30 × 10^−4^** | **1.60 × 10^−2^** |
| **METH-ACETATE-PWY: methanogenesis from acetate** | **All** | **52** | **0.61** | **2.56 × 10^−4^** | **1.60 × 10^−2^** |
| **PWY-6629: superpathway of L-tryptophan biosynthesis** | **All** | **52** | **0.13** | **5.86 × 10^−4^** | **2.34 × 10^−2^** |
| **PWY-5100: pyruvate fermentation to acetate and lactate II** | **All** | **52** | **0.27** | **6.48 × 10^−4^** | **2.34 × 10^−2^** |
| **PWY-7238: sucrose biosynthesis II** | **All** | **52** | **0.23** | **6.70 × 10^−4^** | **2.34 × 10^−2^** |
| **P41-PWY: pyruvate fermentation to acetate and (S)-lactate I** | **All** | **52** | **0.29** | **6.93 × 10^−4^** | **2.34 × 10^−2^** |
| **PWY-7357: thiamine phosphate formation from pyrithiamine and oxythiamine (yeast)** | **All** | **52** | **0.2** | **7.16 × 10^−4^** | **2.34 × 10^−2^** |
| **PWY-6317: D-galactose degradation I (Leloir pathway)** | **All** | **52** | **0.23** | **7.41 × 10^−4^** | **2.34 × 10^−2^** |
| **PWY-8178: pentose phosphate pathway (non-oxidative branch) II** | **All** | **52** | **0.21** | **7.65 × 10^−4^** | **2.34 × 10^−2^** |
| **PWY-5941: glycogen degradation II** | **All** | **52** | **0.23** | **7.91 × 10^−4^** | **2.34 × 10^−2^** |
| **PWY-6823: molybdopterin biosynthesis** | **All** | **52** | **0.33** | **7.91 × 10^−4^** | **2.34 × 10^−2^** |
| **PWY-6572: chondroitin sulfate degradation I (bacterial)** | **All** | **52** | **−0.74** | **8.42 × 10^−4^** | **2.35 × 10^−2^** |
| **GLYCOGENSYNTH-PWY: glycogen biosynthesis I (from ADP-D-Glucose)** | **All** | **52** | **0.24** | **9.32 × 10^−4^** | **2.46 × 10^−2^** |
| **PWY-5973: cis-vaccenate biosynthesis** | **All** | **52** | **−0.23** | **1.03 × 10^−3^** | **2.58 × 10^−2^** |
| **NONOXIPENT-PWY: pentose phosphate pathway (non-oxidative branch) I** | **All** | **52** | **0.24** | **1.17 × 10^−3^** | **2.79 × 10^−2^** |
| **PWY-3001: superpathway of L-isoleucine biosynthesis I** | **All** | **52** | **0.11** | **1.24 × 10^−3^** | **2.82 × 10^−2^** |
| **HSERMETANA-PWY: L-methionine biosynthesis III** | **All** | **52** | **0.18** | **1.41 × 10^−3^** | **2.82 × 10^−2^** |
| **PWY-6122: 5-aminoimidazole ribonucleotide biosynthesis II** | **All** | **52** | **0.1** | **1.41 × 10^−3^** | **2.82 × 10^−2^** |
| **PWY-6277: superpathway of 5-aminoimidazole ribonucleotide biosynthesis** | **All** | **52** | **0.1** | **1.41 × 10^−3^** | **2.82 × 10^−2^** |
| **ARO-PWY: chorismate biosynthesis I** | **All** | **52** | **0.12** | **1.46 × 10^−3^** | **2.82 × 10^−2^** |
| **PWY-8112: factor 420 biosynthesis I (archaea)** | **All** | **52** | **0.37** | **1.66 × 10^−3^** | **3.09 × 10^−2^** |
| **PWY-6609: adenine and adenosine salvage III** | **All** | **52** | **0.09** | **2.05 × 10^−3^** | **3.66 × 10^−2^** |
| **FERMENTATION-PWY: mixed acid fermentation** | **All** | **52** | **0.15** | **2.12 × 10^−3^** | **3.66 × 10^−2^** |
| **ANAGLYCOLYSIS-PWY: glycolysis III (from glucose)** | **All** | **52** | **0.1** | **2.25 × 10^−3^** | **3.71 × 10^−2^** |
| **COBALSYN-PWY: superpathway of adenosylcobalamin salvage from cobinamide I** | **All** | **52** | **0.2** | **2.39 × 10^−3^** | **3.71 × 10^−2^** |
| **PWY-724: superpathway of L-lysine, L-threonine and L-methionine biosynthesis II** | **All** | **52** | **0.1** | **2.39 × 10^−3^** | **3.71 × 10^−2^** |
| **METHANOGENESIS-PWY: methanogenesis from H2 and CO2** | **All** | **52** | **0.39** | **2.53 × 10^−3^** | **3.71 × 10^−2^** |
| **PWY-8113: 3PG-factor 420 biosynthesis** | **All** | **52** | **0.36** | **2.53 × 10^−3^** | **3.71 × 10^−2^** |
| **PWY-5198: factor 420 biosynthesis II (mycobacteria)** | **All** | **52** | **0.35** | **2.58 × 10^−3^** | **3.71 × 10^−2^** |
| **PWY-6936: seleno-amino acid biosynthesis (plants)** | **All** | **52** | **0.3** | **2.69 × 10^−3^** | **3.71 × 10^−2^** |
| **PWY-6386: UDP-N-acetylmuramoyl-pentapeptide biosynthesis II (lysine-containing)** | **All** | **52** | **0.08** | **2.77 × 10^−3^** | **3.71 × 10^−2^** |
| **CALVIN-PWY: Calvin-Benson-Bassham cycle** | **All** | **52** | **0.13** | **2.86 × 10^−3^** | **3.71 × 10^−2^** |
| **PWY-5030: L-histidine degradation III** | **All** | **52** | **−0.38** | **2.95 × 10^−3^** | **3.71 × 10^−2^** |
| **PWY-7663: gondoate biosynthesis (anaerobic)** | **All** | **52** | **−0.24** | **3.03 × 10^−3^** | **3.71 × 10^−2^** |
| **PWY-6168: flavin biosynthesis III (fungi)** | **All** | **52** | **−0.85** | **3.03 × 10^−3^** | **3.71 × 10^−2^** |
| **TRNA-CHARGING-PWY: tRNA charging** | **All** | **52** | **0.07** | **3.22 × 10^−3^** | **3.72 × 10^−2^** |
| **PWY-6163: chorismate biosynthesis from 3-dehydroquinate** | **All** | **52** | **0.1** | **3.31 × 10^−3^** | **3.72 × 10^−2^** |
| **PWY-6892: thiazole component of thiamine diphosphate biosynthesis I** | **All** | **52** | **0.13** | **3.31 × 10^−3^** | **3.72 × 10^−2^** |
| **PWY-5209: methyl-coenzyme M oxidation to CO2** | **All** | **52** | **0.36** | **3.33 × 10^−3^** | **3.72 × 10^−2^** |
| **PWY-6527: stachyose degradation** | **All** | **52** | **0.25** | **3.41 × 10^−3^** | **3.72 × 10^−2^** |
| **PWY-6147: 6-hydroxymethyl-dihydropterin diphosphate biosynthesis I** | **All** | **52** | **−0.29** | **3.51 × 10^−3^** | **3.75 × 10^−2^** |
| **RUMP-PWY: formaldehyde oxidation I** | **All** | **52** | **0.62** | **3.59 × 10^−3^** | **3.75 × 10^−2^** |
| **PWY-4984: urea cycle** | **All** | **52** | **−0.29** | **3.95 × 10^−3^** | **4.04 × 10^−2^** |
| **ARGININE-SYN4-PWY: L-ornithine biosynthesis II** | **All** | **52** | **−0.28** | **4.30 × 10^−3^** | **4.12 × 10^−2^** |
| **HISDEG-PWY: L-histidine degradation I** | **All** | **52** | **−0.29** | **4.30 × 10^−3^** | **4.12 × 10^−2^** |
| **TRPSYN-PWY: L-tryptophan biosynthesis** | **All** | **52** | **0.12** | **4.30 × 10^−3^** | **4.12 × 10^−2^** |
| **PWY-6121: 5-aminoimidazole ribonucleotide biosynthesis I** | **All** | **52** | **0.11** | **4.43 × 10^−3^** | **4.12 × 10^−2^** |
| **PWY-6590: superpathway of Clostridium acetobutylicum acidogenic fermentation** | **All** | **52** | **0.34** | **4.43 × 10^−3^** | **4.12 × 10^−2^** |
| **CENTFERM-PWY: pyruvate fermentation to butanoate** | **All** | **52** | **0.34** | **4.82 × 10^−3^** | **4.40 × 10^−2^** |
| **PWY-7328: superpathway of UDP-glucose-derived O-antigen building blocks biosynthesis** | **All** | **52** | **0.32** | **5.10 × 10^−3^** | **4.58 × 10^−2^** |
| **PWY-6478: GDP-D-glycero-&alpha;-D-manno-heptose biosynthesis** | **All** | **52** | **−0.54** | **5.64 × 10^−3^** | **4.97 × 10^−2^** |
| PWY-7111: pyruvate fermentation to isobutanol (engineered) | All | 52 | 0.19 | 5.87 × 10^−3^ | 5.08 × 10^−2^ |
| PWY-6588: pyruvate fermentation to acetone | All | 52 | 0.95 | 6.29 × 10^−3^ | 5.35 × 10^−2^ |
| VALSYN-PWY: L-valine biosynthesis | All | 52 | 0.09 | 6.56 × 10^−3^ | 5.49 × 10^−2^ |
| PWY-6387: UDP-N-acetylmuramoyl-pentapeptide biosynthesis I (meso-diaminopimelate containing) | All | 52 | 0.07 | 6.93 × 10^−3^ | 5.70 × 10^−2^ |
| FASYN-INITIAL-PWY: superpathway of fatty acid biosynthesis initiation (E. coli) | All | 52 | 0.59 | 7.27 × 10^−3^ | 5.89 × 10^−2^ |
| PWY-7392: taxadiene biosynthesis (engineered) | All | 52 | −0.46 | 9.81 × 10^−3^ | 7.68 × 10^−2^ |
| PWY-5188: tetrapyrrole biosynthesis I (from glutamate) | All | 52 | 0.3 | 1.01 × 10^−2^ | 7.68 × 10^−2^ |
| PWY-6606: guanosine nucleotides degradation II | All | 52 | 0.27 | 1.01 × 10^−2^ | 7.68 × 10^−2^ |
| PWY0-1061: superpathway of L-alanine biosynthesis | All | 52 | 0.45 | 1.01 × 10^−2^ | 7.68 × 10^−2^ |
| PWY-6305: superpathway of putrescine biosynthesis | All | 52 | −0.19 | 1.04 × 10^−2^ | 7.76 × 10^−2^ |
| PWY-6270: isoprene biosynthesis I | All | 52 | 0.11 | 1.06 × 10^−2^ | 7.85 × 10^−2^ |
| PWY-702: L-methionine biosynthesis II | All | 52 | 0.31 | 1.09 × 10^−2^ | 7.92 × 10^−2^ |
| CITRULBIO-PWY: L-citrulline biosynthesis | All | 52 | −0.24 | 1.12 × 10^−2^ | 7.92 × 10^−2^ |
| PWY-6353: purine nucleotides degradation II (aerobic) | All | 52 | 0.25 | 1.12 × 10^−2^ | 7.92 × 10^−2^ |
| GLCMANNANAUT-PWY: superpathway of N-acetylglucosamine, N-acetylmannosamine and N-acetylneuraminate degradation | All | 52 | 0.21 | 1.15 × 10^−2^ | 8.02 × 10^−2^ |
| SALVADEHYPOX-PWY: adenosine nucleotides degradation II | All | 52 | 0.3 | 1.21 × 10^−2^ | 8.33 × 10^−2^ |
| P441-PWY: superpathway of N-acetylneuraminate degradation | All | 52 | 0.36 | 1.24 × 10^−2^ | 8.36 × 10^−2^ |
| PWY-6859: all-trans-farnesol biosynthesis | All | 52 | −0.44 | 1.25 × 10^−2^ | 8.36 × 10^−2^ |
| PWY-5676: acetyl-CoA fermentation to butanoate II | All | 52 | 0.39 | 1.31 × 10^−2^ | 8.53 × 10^−2^ |
| RIBOSYN2-PWY: flavin biosynthesis I (bacteria and plants) | All | 52 | 0.07 | 1.31 × 10^−2^ | 8.53 × 10^−2^ |
| PWY-622: starch biosynthesis | All | 52 | 0.98 | 1.39 × 10^−2^ | 8.87 × 10^−2^ |
| PWY1G-0: mycothiol biosynthesis | All | 52 | 0.25 | 1.43 × 10^−2^ | 8.87 × 10^−2^ |
| GALACTARDEG-PWY: D-galactarate degradation I | All | 52 | −0.77 | 1.43 × 10^−2^ | 8.87 × 10^−2^ |
| GLUCARGALACTSUPER-PWY: superpathway of D-glucarate and D-galactarate degradation | All | 52 | −0.77 | 1.43 × 10^−2^ | 8.87 × 10^−2^ |
| PEPTIDOGLYCANSYN-PWY: peptidoglycan biosynthesis I (meso-diaminopimelate containing) | All | 52 | 0.06 | 1.48 × 10^−2^ | 8.98 × 10^−2^ |
| PWY-6151: S-adenosyl-L-methionine salvage I | All | 52 | 0.09 | 1.48 × 10^−2^ | 8.98 × 10^−2^ |
| PWY-6902: chitin degradation II (Vibrio) | All | 52 | −0.27 | 1.68 × 10^−2^ | 9.71 × 10^−2^ |
| NONMEVIPP-PWY: methylerythritol phosphate pathway I | All | 52 | 0.06 | 1.72 × 10^−2^ | 9.71 × 10^−2^ |
| LACTOSECAT-PWY: lactose and galactose degradation I | All | 52 | 0.44 | 1.77 × 10^−2^ | 9.71 × 10^−2^ |
| PWY-7356: thiamine diphosphate salvage IV (yeast) | All | 52 | 0.36 | 1.77 × 10^−2^ | 9.71 × 10^−2^ |
| PWY-7883: anhydromuropeptides recycling II | All | 52 | −0.47 | 1.78 × 10^−2^ | 9.71 × 10^−2^ |
| COA-PWY-1: superpathway of coenzyme A biosynthesis III (mammals) | All | 52 | 0.09 | 1.81 × 10^−2^ | 9.71 × 10^−2^ |
| PWY-5097: L-lysine biosynthesis VI | All | 52 | 0.06 | 1.81 × 10^−2^ | 9.71 × 10^−2^ |
| PWY-5686: UMP biosynthesis I | All | 52 | 0.07 | 1.81 × 10^−2^ | 9.71 × 10^−2^ |
| PWY-7790: UMP biosynthesis II | All | 52 | 0.07 | 1.81 × 10^−2^ | 9.71 × 10^−2^ |
| PWY-7791: UMP biosynthesis III | All | 52 | 0.07 | 1.81 × 10^−2^ | 9.71 × 10^−2^ |
| PWY-6608: guanosine nucleotides degradation III | All | 52 | 0.2 | 1.86 × 10^−2^ | 9.71 × 10^−2^ |
| PWY-7953: UDP-N-acetylmuramoyl-pentapeptide biosynthesis III (meso-diaminopimelate containing) | All | 52 | 0.05 | 1.86 × 10^−2^ | 9.71 × 10^−2^ |
| PYRIDOXSYN-PWY: pyridoxal 5'-phosphate biosynthesis I | All | 52 | −0.34 | 1.86 × 10^−2^ | 9.71 × 10^−2^ |
| P124-PWY: Bifidobacterium shunt | All | 52 | 0.44 | 1.90 × 10^−2^ | 9.75 × 10^−2^ |
| PWY-7400: L-arginine biosynthesis IV (archaebacteria) | All | 52 | 0.12 | 1.90 × 10^−2^ | 9.75 × 10^−2^ |
| PWY-I9: L-cysteine biosynthesis VI (from L-methionine) | All | 52 | 0.27 | 1.95 × 10^−2^ | 9.89 × 10^−2^ |
| PWY-6385: peptidoglycan biosynthesis III (mycobacteria) | All | 52 | 0.05 | 2.00 × 10^−2^ | 1.00 × 10^−1^ |
| PWY-7209: superpathway of pyrimidine ribonucleosides degradation | All | 52 | 0.49 | 2.09 × 10^−2^ | 1.03 × 10^−1^ |
| PWY-6292: superpathway of L-cysteine biosynthesis (mammalian) | All | 52 | 0.21 | 2.10 × 10^−2^ | 1.03 × 10^−1^ |
| PWY-6749: CMP-legionaminate biosynthesis I | All | 52 | −0.52 | 2.14 × 10^−2^ | 1.04 × 10^−1^ |
| PWY-6160: 3-dehydroquinate biosynthesis II (archaea) | All | 52 | 0.26 | 2.38 × 10^−2^ | 1.14 × 10^−1^ |
| PWY-6165: chorismate biosynthesis II (archaea) | All | 52 | 0.33 | 2.38 × 10^−2^ | 1.14 × 10^−1^ |
| GLUCONEO-PWY: gluconeogenesis I | All | 52 | 0.11 | 2.42 × 10^−2^ | 1.15 × 10^−1^ |
| ILEUSYN-PWY: L-isoleucine biosynthesis I (from threonine) | All | 52 | 0.09 | 2.48 × 10^−2^ | 1.16 × 10^−1^ |
| PWY-4041: &gamma;-glutamyl cycle | All | 52 | 0.46 | 2.60 × 10^−2^ | 1.21 × 10^−1^ |
| PWY-6630: superpathway of L-tyrosine biosynthesis | All | 52 | 0.26 | 2.66 × 10^−2^ | 1.21 × 10^−1^ |
| PWY-1861: formaldehyde assimilation II (assimilatory RuMP Cycle) | All | 52 | 0.34 | 2.66 × 10^−2^ | 1.21 × 10^−1^ |
| P461-PWY: hexitol fermentation to lactate, formate, ethanol and acetate | All | 52 | 0.42 | 2.79 × 10^−2^ | 1.26 × 10^−1^ |
| PWY-7115: C4 photosynthetic carbon assimilation cycle, NAD-ME type | All | 52 | 0.4 | 2.92 × 10^−2^ | 1.31 × 10^−1^ |
| PWY-6126: superpathway of adenosine nucleotides de novo biosynthesis II | All | 52 | −0.15 | 3.13 × 10^−2^ | 1.39 × 10^−1^ |
| THISYNARA-PWY: superpathway of thiamine diphosphate biosynthesis III (eukaryotes) | All | 52 | 0.12 | 3.35 × 10^−2^ | 1.47 × 10^−1^ |
| PWY-6396: superpathway of 2,3-butanediol biosynthesis | All | 52 | 0.21 | 3.46 × 10^−2^ | 1.51 × 10^−1^ |
| PWY-6700: queuosine biosynthesis I (de novo) | All | 52 | 0.06 | 3.58 × 10^−2^ | 1.55 × 10^−1^ |
| P621-PWY: nylon-6 oligomer degradation | All | 52 | 0.29 | 3.60 × 10^−2^ | 1.55 × 10^−1^ |
| COA-PWY: coenzyme A biosynthesis I (prokaryotic) | All | 52 | 0.08 | 3.74 × 10^−2^ | 1.58 × 10^−1^ |
| PWY-7228: superpathway of guanosine nucleotides de novo biosynthesis I | All | 52 | −0.16 | 3.74 × 10^−2^ | 1.58 × 10^−1^ |
| BRANCHED-CHAIN-AA-SYN-PWY: superpathway of branched chain amino acid biosynthesis | All | 52 | 0.08 | 3.91 × 10^−2^ | 1.61 × 10^−1^ |
| PWY-6125: superpathway of guanosine nucleotides de novo biosynthesis II | All | 52 | −0.18 | 3.91 × 10^−2^ | 1.61 × 10^−1^ |
| PWY-7851: coenzyme A biosynthesis II (eukaryotic) | All | 52 | 0.08 | 3.91 × 10^−2^ | 1.61 × 10^−1^ |
| ASPASN-PWY: superpathway of L-aspartate and L-asparagine biosynthesis | All | 52 | −0.09 | 4.09 × 10^−2^ | 1.66 × 10^−1^ |
| P164-PWY: purine nucleobases degradation I (anaerobic) | All | 52 | 0.22 | 4.09 × 10^−2^ | 1.66 × 10^−1^ |
| PWY-7220: adenosine deoxyribonucleotides de novo biosynthesis II | All | 52 | −0.22 | 4.27 × 10^−2^ | 1.70 × 10^−1^ |
| PWY-7222: guanosine deoxyribonucleotides de novo biosynthesis II | All | 52 | −0.22 | 4.27 × 10^−2^ | 1.70 × 10^−1^ |
| PWY-7229: superpathway of adenosine nucleotides de novo biosynthesis I | All | 52 | −0.11 | 4.37 × 10^−2^ | 1.73 × 10^−1^ |
| PWY-1042: glycolysis IV | All | 52 | 0.07 | 4.46 × 10^−2^ | 1.75 × 10^−1^ |
| PHOSLIPSYN-PWY: superpathway of phospholipid biosynthesis I (bacteria) | All | 52 | −0.15 | 4.56 × 10^−2^ | 1.77 × 10^−1^ |
| PWY-6470: peptidoglycan biosynthesis V (&beta;-lactam resistance) | All | 52 | 0.18 | 4.76 × 10^−2^ | 1.84 × 10^−1^ |
| POLYISOPRENSYN-PWY: polyisoprenoid biosynthesis (E. coli) | All | 52 | −0.25 | 4.86 × 10^−2^ | 1.85 × 10^−1^ |
| PWY-5103: L-isoleucine biosynthesis III | All | 52 | 0.09 | 4.86 × 10^−2^ | 1.85 × 10^−1^ |
| PWY4FS-7: phosphatidylglycerol biosynthesis I (plastidic) | All | 52 | −0.19 | 4.97 × 10^−2^ | 1.85 × 10^−1^ |
| PWY4FS-8: phosphatidylglycerol biosynthesis II (non-plastidic) | All | 52 | −0.19 | 4.97 × 10^−2^ | 1.85 × 10^−1^ |
| SER-GLYSYN-PWY: superpathway of L-serine and glycine biosynthesis I | All | 52 | 0.1 | 4.97 × 10^−2^ | 1.85 × 10^−1^ |

The table summarizes microbial metabolic pathways that changed significantly between the pre- and post-intervention time points based on paired Wilcoxon signed-rank tests. The Group column indicates the analyzed participant group, N pairs denotes the number of paired samples included in the analysis, log2 Fold Change represents the direction and magnitude of the pathway abundance change following the intervention, Wilcoxon p indicates the nominal p-value, and FDR-adjusted p represents the false discovery rate-adjusted p-value using the Benjamini–Hochberg procedure. Positive log2 fold-change values indicate increased pathway abundance after the intervention, whereas negative values indicate decreased abundance. **Green rows indicate pathways that remained statistically significant after false discovery rate (FDR) correction.**

**References**

1.	Tanaka H, Monahan KD, Seals DR. Age-predicted maximal heart rate revisited. *Journal of the American College of Cardiology.* 2001;37(1):153-156.

2.	Jokai M, Torma F, McGreevy KM, et al. DNA methylation clock DNAmFitAge shows regular exercise is associated with slower aging and systemic adaptation. *GeroScience.* 2023;45(5):2805-2817.

3.	Leung JL, Lee GT, Lam YH, Chan RC, Wu JY. The use of the Digit Span Test in screening for cognitive impairment in acute medical inpatients. *Int Psychogeriatr.* 2011;23(10):1569-1574.

4.	Woods DL, Kishiyamaa MM, Lund EW, et al. Improving digit span assessment of short-term verbal memory. *J Clin Exp Neuropsychol.* 2011;33(1):101-111.

5.	Horvath S, Oshima J, Martin GM, et al. Epigenetic clock for skin and blood cells applied to Hutchinson Gilford Progeria Syndrome and ex vivo studies. *Aging (Albany NY).* 2018;10(7):1758-1775.

6.	McGreevy KM, Radak Z, Torma F, et al. DNAmFitAge: biological age indicator incorporating physical fitness. *Aging (Albany NY).* 2023;15(10):3904-3938.

7.	Lu AT, Binder AM, Zhang J, et al. DNA methylation GrimAge version 2. *Aging (Albany NY).* 2022;14(23):9484-9549.

8.	Lu AT, Quach A, Wilson JG, et al. DNA methylation GrimAge strongly predicts lifespan and healthspan. *Aging (Albany NY).* 2019;11(2):303-327.

9.	Hannum G, Guinney J, Zhao L, et al. Genome-wide methylation profiles reveal quantitative views of human aging rates. *Mol Cell.* 2013;49(2):359-367.

10.	Belsky DW, Caspi A, Corcoran DL, et al. DunedinPACE, a DNA methylation biomarker of the pace of aging. *Elife.* 2022;11.

11.	Nasser S, Keringer J, Gu Y, et al. Physical Fitness Is Negatively Associated With DNA Methylation-Based Risk of Aging-Related Diseases. *Aging cell.* 2026;25(4):e70467.

12.	Lu AT, Seeboth A, Tsai PC, et al. DNA methylation-based estimator of telomere length. *Aging (Albany NY).* 2019;11(16):5895-5923.

13.	Higgins-Chen AT, Thrush KL, Wang Y, et al. A computational solution for bolstering reliability of epigenetic clocks: Implications for clinical trials and longitudinal tracking. *Nat Aging.* 2022;2(7):644-661.

14.	Ewels P, Magnusson M, Lundin S, Kaller M. MultiQC: summarize analysis results for multiple tools and samples in a single report. *Bioinformatics.* 2016;32(19):3047-3048.

15.	Chen S, Zhou Y, Chen Y, Gu J. fastp: an ultra-fast all-in-one FASTQ preprocessor. *Bioinformatics.* 2018;34(17):i884-i890.

16.	Langmead B, Salzberg SL. Fast gapped-read alignment with Bowtie 2. *Nat Methods.* 2012;9(4):357-359.

17.	Blanco-Miguez A, Beghini F, Cumbo F, et al. Extending and improving metagenomic taxonomic profiling with uncharacterized species using MetaPhlAn 4. *Nat Biotechnol.* 2023;41(11):1633-1644.

18.	Beghini F, McIver LJ, Blanco-Miguez A, et al. Integrating taxonomic, functional, and strain-level profiling of diverse microbial communities with bioBakery 3. *Elife.* 2021;10.

19.	Suzek BE, Wang Y, Huang H, McGarvey PB, Wu CH, UniProt C. UniRef clusters: a comprehensive and scalable alternative for improving sequence similarity searches. *Bioinformatics.* 2015;31(6):926-932.

20.	Simpson. EH. Measurement of diversity. *Nature.* 1949;163:688.

21.	Javier Palarea-Albaladejo JAM-F. zCompositions—R package for multivariate imputation of left-censored data under a compositional approach. *Chemometrics and Intelligent Laboratory Systems,.* 2015.;143:85–96.

22.	Josep A. Martín-Fernández KH, Matthias Templ, Peter Filzmoser, and Javier Palarea-Albaladejo. Bayesian-multiplicative treatment of count zeros in compositional data sets. *Statistical Modelling.* 2015;15(2):134–158.

23.	Aitchison. J. *The Statistical Analysis of Compositional Data.* London: Chapman and Hall; 1986.

24.	Aton M, McDonald D, Canardo Alastuey J, et al. Scikit-bio: a fundamental Python library for biological omic data analysis. *Nat Methods.* 2026;23(2):274-276.

25.	Valentin Todorov PF. An Object-Oriented Framework for Robust Multivariate Analysis. *Journal of Statistical Software.* 2009;32(3):1-47.

26.	Manuel Koller WAS. Sharpening Wald-type inference in robust regression for small samples. *Computational Statistics & Data Analysis.* 2011;55(8):2504-2515.

27.	Torma F, Kerepesi C, Jokai M, et al. Alterations of the gut microbiome are associated with epigenetic age acceleration and physical fitness. *Aging cell.* 2024;23(4):e14101.

28.	Swain DP, Leutholtz BC, King ME, Haas LA, Branch JD. Relationship between % heart rate reserve and % VO2 reserve in treadmill exercise. *Med Sci Sports Exerc.* 1998;30(2):318-321.

29.	Teglas T, Torma F, Bori Z, et al. Epigenetic insights of Olympic champions: nuclear and mitochondrial DNA methylation and regulators of aging. *GeroScience.* 2026.
